# Projecting excess mortality due to infectious diseases during a crisis: methods and application to the Gaza Strip, 2023-2024

**DOI:** 10.1101/2025.06.11.25329427

**Authors:** Francesco Checchi, Gregory Barnsley, Zeina Jamaluddine, Hanan Abukmail, Zhixi Chen, Paul B Spiegel, Fiona Majorin

**Affiliations:** Faculty of Epidemiology and Population Health, London School of Hygiene and Tropical Medicine, Keppel St, London; Center for Humanitarian Health, Johns Hopkins Bloomberg School of Public Health, Baltimore, MD, United States of America

**Keywords:** infection, epidemic, endemic, humanitarian, crisis, war, Gaza, model, projection

## Abstract

**Background:** Excess mortality due to infectious diseases is a key consequence of crises due to armed conflicts and natural disasters, but is measured retrospectively if at all. The Israeli operation in the Gaza Strip has exposed the population to known risk factors for increased infectious disease burden (overcrowding, malnutrition, inadequate water, sanitation and hygiene, disrupted health services). We developed a novel method for forward-projecting mortality due to both endemic and epidemic-prone infections in Gaza between 7 October 2023 and 6 August 2024 (ten months).

**Methods:** After defining alternative crisis scenarios, we used expert elicitation to quantify the probability of specific epidemics and increases in transmissibility and case-fatality. We analysed past vaccination coverage and natural exposure information to estimate population susceptibility. For endemic infections, we took pre-war deaths, adjusted for COVID-19, as the counterfactual baseline, multiplied by the above parameters to compute projections. For epidemics, we projected deaths by simulating a susceptible-exposed-infected-recovered process. All analyses were by disease and age and implemented stochastically to propagate parameter uncertainty.

**Results:** Over the total period, we estimated that about 4000 endemic infection deaths would occur under a status quo scenario, ranging from 3400 to 4600 under ceasefire and escalation scenarios and comprising mainly COVID-19, influenza and pneumococcal disease. For epidemics, we projected 8500 deaths under the status quo but with a wide 95% confidence interval (0 to 100,300, rising to 128,900 under military escalation), reflecting high uncertainty about whether epidemics would indeed occur; cholera, measles and polio were the most likely epidemic threats.

**Conclusion:** The model generated actionable projections for the Gaza Strip under a single framework covering all infectious diseases of public health interest and accounting for the effects of crisis on immunity, transmissibility and case-fatality. The method needs to be improved, e.g. by explicitly featuring the effect of malnutrition and strengthening expert elicitation techniques, before it can be generalised to other crises.

## Background

Accurate mortality estimates provide a quantifiable measure of a crisis’ impact on population health, both directly through injuries or indirectly through the disruption of essential services and increased exposure to risk factors [1]. These estimates are essential for grading the severity of a crisis, adjusting relief operations, and advocating for necessary assistance or compliance with international humanitarian law [2]. Excess mortality, i.e. deaths above and beyond the counterfactual level that would have occurred in the absence of a crisis [1], is of particular relevance.

While retrospective (e.g. through surveys) or real-time (prospective surveillance) mortality estimates are common in crisis settings, forward-projections, to our knowledge, have not been attempted. Looking into the future could usefully guide humanitarian response and local public health initiatives and provide a basis for advocacy.

Since 7 October 2023’s Hamas attacks, Israel’s large-scale military operations in the Gaza Strip, including aerial bombing and ground offensives, have resulted in a severe public health crisis. As of 22 April 2025, 51,266 people had been reported killed and 116,991 injured [3], with injury deaths likely underestimated by about 40% at least until June 2024 [4]. About 85% of Gazans were displaced or in need of emergency shelter, with widespread damage to water, sanitation and hygiene (WASH) infrastructure [5] resulting in poor sanitation conditions and water provision below Sphere standards for most. Both outpatient and inpatient health services were heavily disrupted [6] and subject to military attacks or forced closures [7]. Fluctuating levels of food provision had made most of the population food-insecure, though malnutrition levels had remained moderate at least during 2024 [8]. Despite these highly prevalent risk factors, empirical estimates of deaths indirectly attributable to the war have not yet been published to our knowledge.

In order to inform humanitarian and decision-makers working on the Gaza crisis, we estimated the potential public health impact of the crisis under different possible scenarios (www.gaza-projections.org) using a suite of mechanistic models. These projections covered a six-month period from 7 February to 6 August 2024 while also estimating deaths since 7 October 2023. We stratified mortality into five cause-specific ‘modules’: traumatic injuries, infectious diseases, maternal and neonatal deaths and stillbirths, and non-communicable diseases (NCDs), with malnutrition as an underlying cause. Here, we present in-depth methods and findings for the infection module, discuss limitations and outline possible improvements to the approach so as to enable its application in other crises.

## Methods

### Study scope

#### Population and period

The study covered the entire population of the Gaza Strip over the period from 7 October 2023 to 6 August 2024, with the period up to 6 February 2024 (four months) henceforth referred to as ‘retrospective’, insofar as our estimates were, to the extent possible, based on observed data and drawn up at the end of this period; by contrast, the period 7 February to 6 August (six months) was subject to forward projection under alternative scenarios (see below). We assumed a constant population size throughout the analysis period, estimated using census and United Nations projections (Supplementary Information).

#### Diseases analysed

We included both *endemic* (defined as a relatively constant secular trend in terms of the number of infections per year, with or without a seasonal peak) and *epidemic-prone* (defined as occurrence of at least three generations of uninterrupted transmission of the pathogen within the general population, excluding instances of sustained transmission within specific groups such as hospitalised patients) infections within our analysis (Table 1).

**Table 1.** Infectious diseases (pathogens) included within the scope of projections.

| Included in Gaza routine vaccination schedule [9]? | Main route of transmission | Epidemic-prone infectious diseases (pathogens) | Endemic infectious diseases (pathogens) (W = highly seasonal) |
| --- | --- | --- | --- |
| yes | Airborne-droplet | Diphtheria (*Corynebacterium diphtheriae*)<br>Pertussis (*Bordetella pertussis*)<br>Measles (measles virus) | *Haemophilus influenzae* type b disease (*Haemophilus influenzae* type B)<br>Pneumococcal disease (*Streptococcus pneumoniae*)<br>COVID-19‡ (SARS-CoV-2 virus) |
|  | Faecal-oral | Polio (wild-type poliovirus type 1 and 3) | Rotavirus disease (rotavirus) (W) |
| no | Airborne-droplet | Meningococcal meningitis (*Neisseria meningitidis* types A, C, W or Y)† | Influenza†, para-influenza (human influenza, para-influenza viruses) (W)<br>Respiratory syncytial virus disease, RSV (respiratory syncytial virus) (W) |
|  | Faecal-oral | Bacterial dysentery (*Shigella dysenteriae* type 1)<br>Cholera (*Vibrio cholerae*)<br>Hepatitis A (hepatitis A virus)<br>Hepatitis E (hepatitis E virus)<br>Polio (vaccine-derived poliovirus type 2)<br>Typhoid fever (*Salmonella typhi*) | Bacterial gastroenteritis other than epidemic-prone causes (*Salmonella typhimurium*, *Escherichia coli*, *Clostridium difficile*, *Campylobacter jejuni*, *Shigella* spp.)<br>Viral gastroenteritis other than rotavirus (adenovirus, astrovirus, norovirus) (W) |

† Vaccination only recommended for vulnerable groups. ‡ Unclear status of vaccination coverage post-2022, with low uptake up to 2022.

Tuberculosis, HIV, sexually transmitted infections, hepatitis B and C, mumps, rubella, rabies, chickenpox, tetanus, neonatal infections and vector-borne infections were excluded from analysis due to their low case fatality ratio (CFR), low baseline burden according to pre-crisis Ministry of Health epidemiological data [10] and/or being covered by the maternal and neonatal deaths and stillbirths (neonatal infections) or traumatic injury (infected wounds) modules of the wider project. We also excluded antimicrobial resistance (AMR) due to the difficulty in separating deaths due to AMR from those due to infection as a primary cause. Concerns about the spread of AMR due to the war in Gaza have nonetheless been expressed [11].

#### Projection scenarios

After consulting with diplomatic and humanitarian experts familiar with the crisis, we developed three possible scenarios for how the crisis would progress over the projection period:

1. *Ceasefire* (reasonable-best): A long-term or permanent cessation of military activity, with increased humanitarian access and mobility;
2. *Status quo* (likely): Continuation of conditions experienced hitherto (specifically, the period 15 October 2023 to 15 January 2024);
3. *Escalation* (reasonable-worst): Further intensification of military operations and resulting risk factors.

For each projection, we described characteristics such as occurrence and duration of any pauses/ceasefires, intensity and typology of military activity, etc. Assumptions were then made about the values of key factors (water, sanitation and hygiene; nutrition; shelter) and health system disruptions (vaccination, access to rehydration and antimicrobials, etc.) that would have proximally modulated infection transmissibility and/or case-fatality, based on a causal framework (see Supplementary Information, Figure S1 and [1]). Further details on the scenarios are provided in the Supplementary Information.

### Analytic framework

#### Epidemic-prone infections model

All epidemic-prone pathogens under consideration were modelled using a Susceptible - Exposed - Infectious - Recovered (SEIR) framework [12]. The term ‘exposed’ refers to people infected but not yet infectious, and ‘recovered’ comprises those who are immune due to natural or vaccine-induced exposure. Transitions among compartments are governed by 𝜆, the force of infection (incidence acting on susceptibles), 𝜌, the rate of transition out of the pre-infectious state (where ρ^−1^ is the mean pre-infectious period) and 𝛾, the rate of transition out of the infectious state (where 𝛾^−1^ is the mean duration of infectiousness). Given the short timeframe of the projection period, we assumed zero waning of immunity back into the susceptible compartment.

We modelled age-specific deaths 𝐷_𝑎_ attributable to epidemics of each disease 𝑢 as realisations of a SEIR process, conditional on the binomial probability of an epidemic occurring during the projection period 𝑇 :

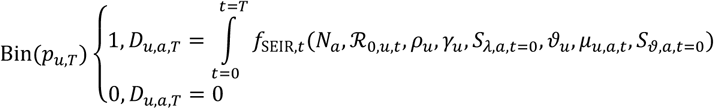

where 𝑝_𝑢,𝑇_ is the probability of an epidemic during the period; 𝑁_𝑎_ is age-specific population; ℛ_0,𝑢,𝑡_ is the basic reproduction number and is allowed to vary over time; 𝑆_𝜆,𝑎,𝑡=0_ and 𝑆_𝜗,𝑎,𝑡=0_ are the number of people susceptible to infection and disease, respectively; 𝜗_𝑢_ is the proportion of infections that result in symptomatic disease; and 𝜇_𝑢,𝑎,𝑡_ is the CFR among symptomatic cases, also time-varying.

We computed deaths over each time step by multiplying SEIR-modelled incident infections by the proportion symptomatic and by CFR, corrected for the time-dependent probability of being immune to severe disease, conditional on not being immune to infection:

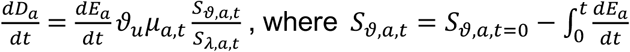

For each pathogen 𝑢, we implemented an age-structured SEIR model with the following age groups: 0 months old (mo), 1 to 11mo, 12 to 59mo, 5 to 14 years old (yo), 15 to 19yo, and thereafter ten-year age groups with the oldest age group being ≥ 80yo. We ignored births, deaths and aging due to the short analysis timeframe. The force of infection 𝜆 was computed assuming transmissibility ℛ_0_⁄𝛾 (where ℛ_0_ is the basic reproduction number) and a heterogeneous age-specific contact structure, for which we used the R socialmixr [13] package to upload and prepare a contact matrix previously estimated among a population of camp-based internally displaced persons in Somaliland, which to our knowledge constitutes the only available dataset of social contact structure among displaced people [14] (Figure S2). Lastly, we initialised the 𝐼_𝑎,𝑡=0_ compartment as 1 per million population per age group, with 𝐸_𝑎,𝑡=0_ = 0 and 𝑅_𝑎,𝑡=0_ = 𝑁_𝑎_ − 𝑆_𝜆,𝑎,𝑡=0_ − 𝐼_𝑎,𝑡=0_.

We used the R package epidemics [15] to implement multiple runs of each disease-specific SEIR model over daily time increments, and compute cumulative infections at the end of the projection period. For each simulation, we propagated uncertainty as follows: (i) we sampled from an empirical distribution of 𝑝_𝑢_ and drew a random binomial realisation from the sampled 𝑝_𝑢_ value to decide whether an epidemic would in fact take place during the period; (ii) conditional on an epidemic taking place, we selected a random epidemic starting day within the projection period; (iii) we sampled from empirical distributions of ℛ_0_ and 𝜇_𝑎_ (the age-specific CFR); and from a uniform distribution of the range of 𝜗_𝑢_, the proportion of symptomatic infections. We extracted estimates of 𝜌, 𝛾 and 𝜗_𝑢_from the literature (Table 2). Sources of values/distributions for other (all crisis-specific) parameters are described below.

**Table 2.**
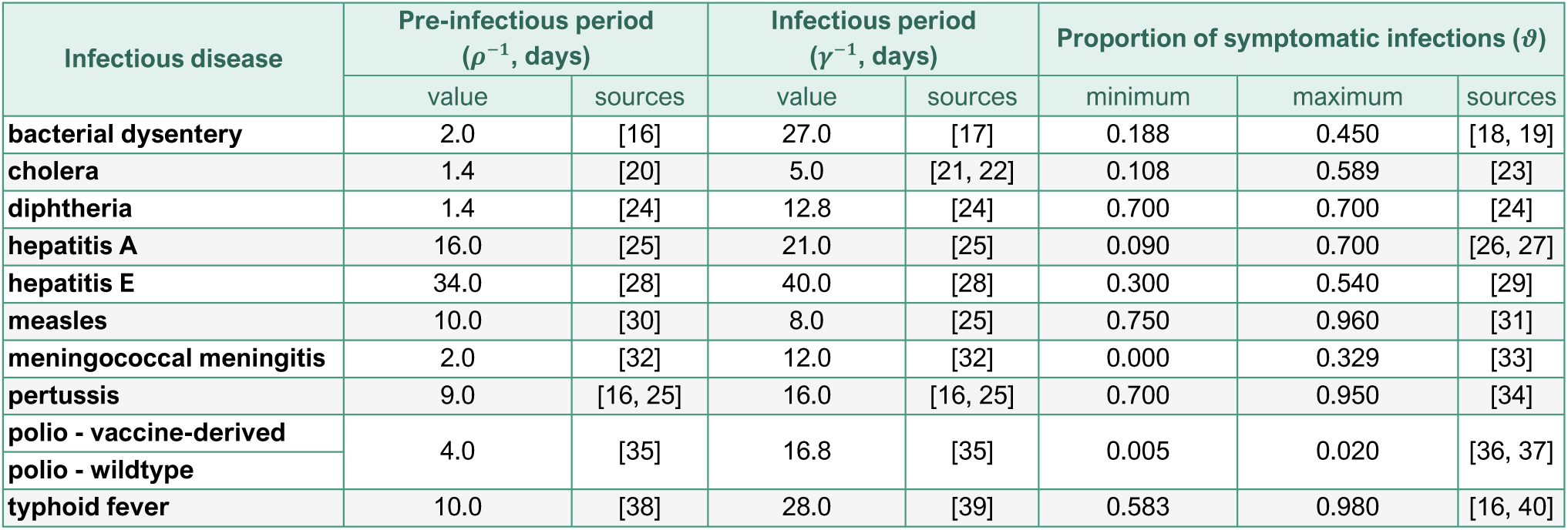
Estimates of the pre-infectious and infectious period and of the proportion of symptomatic infections, by infectious pathogen.

| Infectious disease | Pre-infectious period ($\rho^{-1}$, days) | | Infectious period ($\gamma^{-1}$, days) | | Proportion of symptomatic infections ($\vartheta$) | | |
| --- | --- | --- | --- | --- | --- | --- | --- |
|  | value | sources | value | sources | minimum | maximum | sources |
| bacterial dysentery | 2.0 | [16] | 27.0 | [17] | 0.188 | 0.450 | [18, 19] |
| cholera | 1.4 | [20] | 5.0 | [21, 22] | 0.108 | 0.589 | [23] |
| diphtheria | 1.4 | [24] | 12.8 | [24] | 0.700 | 0.700 | [24] |
| hepatitis A | 16.0 | [25] | 21.0 | [25] | 0.090 | 0.700 | [26, 27] |
| hepatitis E | 34.0 | [28] | 40.0 | [28] | 0.300 | 0.540 | [29] |
| measles | 10.0 | [30] | 8.0 | [25] | 0.750 | 0.960 | [31] |
| meningococcal meningitis | 2.0 | [32] | 12.0 | [32] | 0.000 | 0.329 | [33] |
| pertussis | 9.0 | [16, 25] | 16.0 | [16, 25] | 0.700 | 0.950 | [34] |
| polio - vaccine-derived | 4.0 | [35] | 16.8 | [35] | 0.005 | 0.020 | [36, 37] |
| polio - wildtype |  |  |  |  |  |  |  |
| typhoid fever | 10.0 | [38] | 28.0 | [39] | 0.583 | 0.980 | [16, 40] |

We implemented 10,000 simulation runs and computed the mean and 95% percentile interval of cumulative deaths. All epidemic-attributable deaths are counted as excess mortality, due to the absence of major epidemics in Gaza over the decade prior to the war. As no epidemics were confirmed in Gaza during the ‘retrospective’ period (up to 6 February 2024), we only ran the model for the projection period and assumed zero epidemic-attributable mortality earlier in the crisis.

### Endemic infections model

For each endemic infection 𝑢 and age group 𝑎, we estimated mortality during the retrospective and projection periods as a function of a counterfactual baseline drawn from pre-war data (see below), multiplied by relative risks of transmission 𝜑_𝜆,𝑢,𝑡_ and case-fatality 𝜑_𝜇,𝑢,𝑡_, with adjustment for the seasonal pattern of incidence 𝜔_𝑢,𝑡_, as follows:

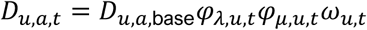

Specifically, 𝜑_𝜆,𝑢,𝑡_ and 𝜑_𝜇,𝑢,𝑡_ are the ratios of, respectively, ℛ_0,𝑢_ and CFR during the crisis to ℛ_0,𝑢_ and CFR under the counterfactual baseline, while 𝜔_𝑢,𝑡_ is the ratio of cases expected during the calendar month that 𝑡 falls within to the mean number of monthly cases over a typical calendar year. Sources of values for these parameters are described below. Notably, we decoupled susceptibility from 𝜑_𝜆,𝑢,𝑡_ and 𝜑_𝜇,𝑢,𝑡_, as we assumed that, over the relatively short analysis period, (i) for infections included in the pre-war vaccination programme (pneumococcus, Hib, rotavirus) herd immunity built up over the past years of very high vaccination coverage (Figure S3) would largely have protected the most vulnerable; while (ii) for infections not included in the routine vaccination programme (COVID-19, influenza and para-influenza, RSV, other bacterial gastroenteritis) crisis-emergent factors (e.g. faster immunological waning due to malnutrition) would have had a negligible effect on susceptibility.

We implemented 10,000 runs of the endemic infections model, each time sampling from the prediction of the negative-binomial model of baseline (counterfactual) deaths and the distributions of 𝜑_𝜆,𝑢,𝑡_ and 𝜑_𝜇,𝑢,𝑡_. In each run, we calculated excess deaths as 𝐷_𝑢,𝑎,𝑡_ − 𝐷_𝑢,𝑎,base_. We computed the mean and 95% percentile intervals of run outputs.

### Model parameterisation

#### Baseline and crisis-period susceptibility estimates

For infections included in Gaza’s routine vaccination schedule, we developed an age-structured dynamic cohort model to track population susceptibility over time based on vaccination coverage, natural exposure to infection, vaccine effectiveness estimates and immunological waning. Age strata were monthly until 23mo, yearly until 6yo, 6 to 9yo, 10 to 14yo and thereafter as per the epidemic model (above).

As represented in Figure 1, children are born into either the 𝑀 (maternally immune) or 𝑆 (susceptible compartment) based on susceptibility in the reproductive age group and a birth rate 𝑏. Children lose their maternal immunity at a rate 𝑚. They also receive their routine vaccinations as they age, moving to compartment 𝑉_𝜆_ (people with vaccine-induced immunity against infection) based on the product of 𝑐, vaccination coverage, and 𝑓_𝜆_, vaccine effectiveness against infection; vaccination is non-specific, i.e. immune children are also vaccinated. Vaccination may also occur later in life, as booster doses. All susceptible compartments experience a force of infection 𝜆. We also track compartment 𝑉_𝜎_ (not shown; people with vaccine-induced immunity against disease, but not infection) as the product of 𝑐 and 𝑓_𝜎_, vaccine effectiveness against severe disease.

**Figure 1.**
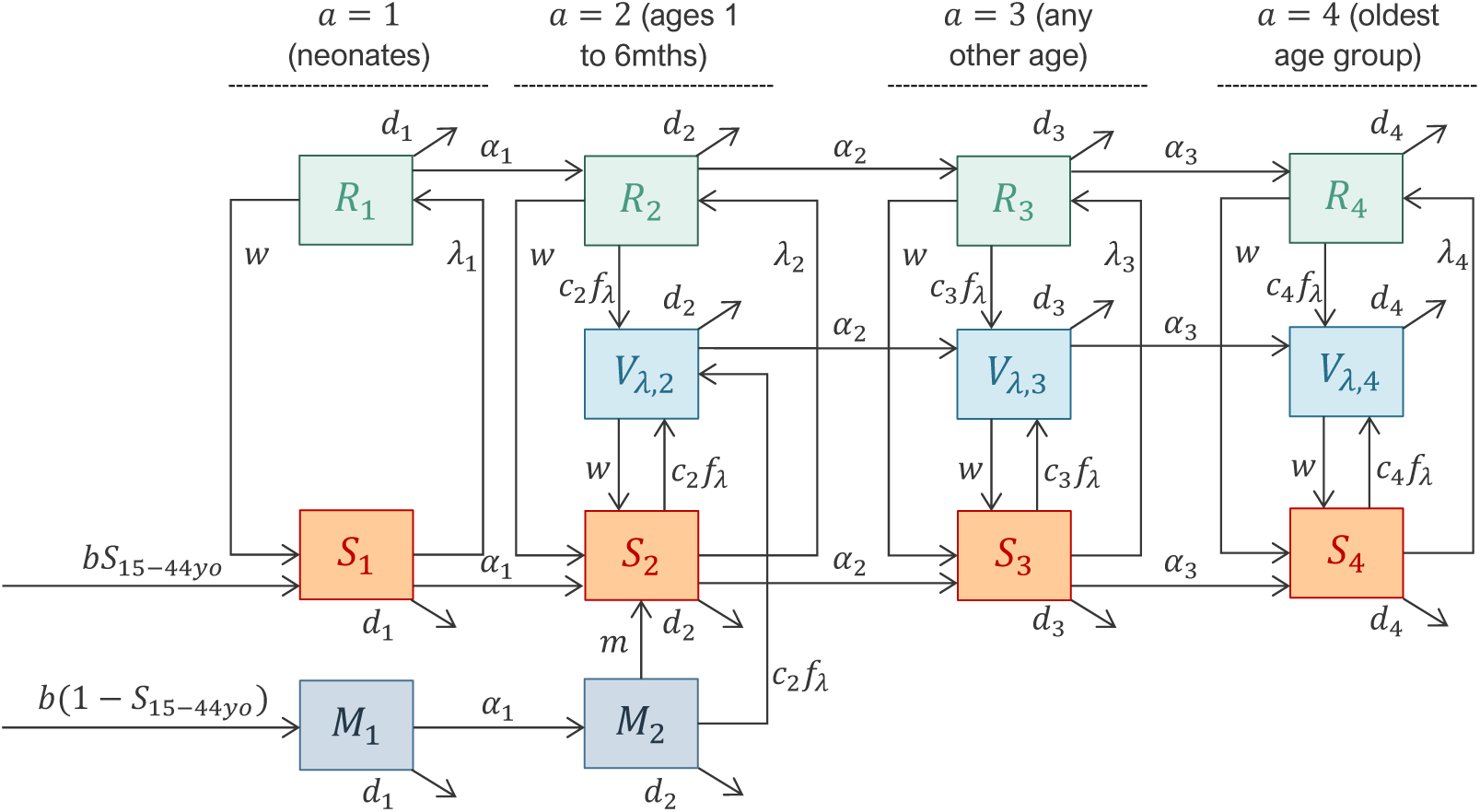
Diagram showing states and flows in the population immunity model, for a single pathogen. For brevity, only a few age groups (numeric subscripts) are shown and vaccine derived protection against disease but not infection is not shown as it is treated identically to full vaccine protection but must be explicitly tracked for the modelling.

We made simplifying assumptions: (i) waning time is negative-exponentially distributed with rate 𝑤 = 𝑇_waning_^−1^, the inverse of the mean duration of functional protection, and equal for protection against infection and against disease, and for natural and vaccine-acquired immunity; (ii) maternal immunity protection lasts 𝑇_M_ = 6 months, roughly corresponding to the observed mean duration of exclusive breastfeeding in Gaza, and estimates of this parameter for different vaccines and infections; (iii) vaccines have constant effectiveness by age and confer ‘all-or-nothing’ protection; (iv) vaccinations occur as a proportion of those ageing into the relevant age group per the theoretical dosage schedules (Figure S3); and (v) people acquire vaccine-derived immunity when they complete the full age-appropriate priming regimen, with subsequent booster doses undoing any waning. Table S6 specifies differential equations quantifying the evolution of each model compartment. Vaccine effectiveness and waning estimates from the literature are summarised in Table S7. Table S8 summarises baseline susceptibility assumptions for infections not included in Gaza’s vaccination programme.

We simulated the cohort from January 2000 to the end of the projection period, using historical (Figure S3) and scenario-specified (Table S3) vaccination coverage, crude birth and death rates, the latter distributed across age groups based on Ministry of Health mortality statistics for 2019-2022 [41–44]. As no cases of polio and diphtheria were reported during this historical period, we assumed there was no naturally acquired immunity to these diseases. For some infections we were able to apply Ministry of Health surveillance reports to estimate annual 𝜆 (zero reported cases for polio and diphtheria; sporadic outbreaks of measles and Hib). For pertussis, pneumococcus and rotavirus, owing to lack of specific surveillance we assumed an annual base 𝜆 of 500 per 1000 under age 24mo, drawing on studies from Gaza or elsewhere [45, 46]. This 𝜆 was allowed to vary over time as immunity changed and vaccines were introduced.

#### Counterfactual (baseline) mortality by age, sex and season

We fitted a negative binomial model to the reported yearly number of deaths due to infections during 2016-2022, offset by ln 𝑁 and with the proportion of deaths due to COVID-19 as the single explanatory variable. The model had reasonable fit to the data (Figure 2), and we used it to forecast a counterfactual value of all-age infection deaths in 2023 and 2024, setting the percent due to COVID-19 at 30%, as per COVID-19 proportional mortality in England and Wales during the first half of 2023 [47].

**Figure 2.**
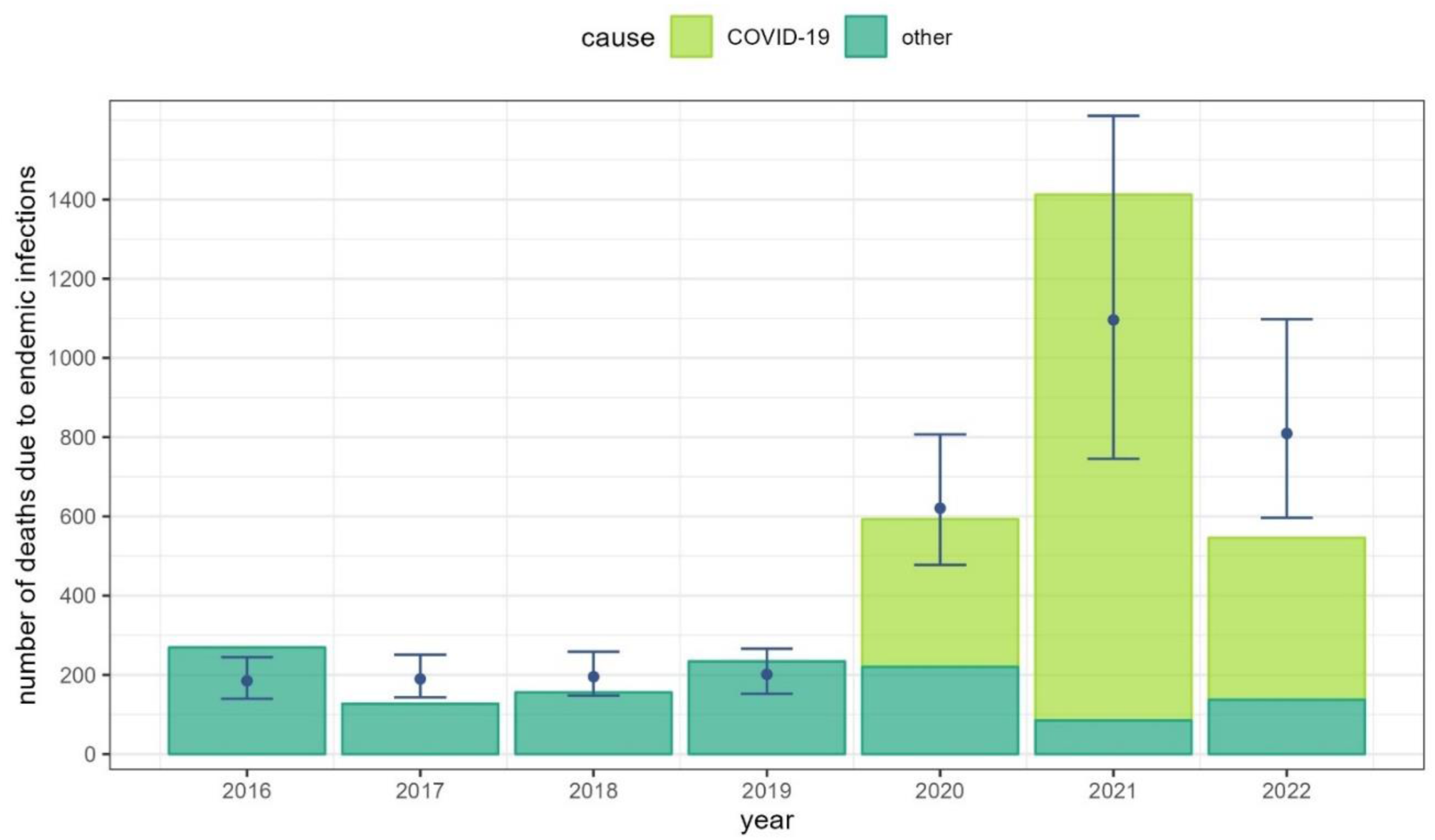
Number of deaths due to infectious diseases per year, by cause (COVID-19 or other). Source: Ministry of Health, Gaza [10]. The blue dots and error bars denote, respectively, the mean and 95%CI of a negative binomial model fit to the data.

As no epidemics were reported during 2016-2022, we assumed that none would have occurred during the analysis period and set counterfactual epidemic mortality to zero. We thus attributed the total counterfactual deaths to specific endemic infections based on available pre-war data from the Gaza Strip, or, if unavailable, the West Bank. In Gaza in 2017 infection deaths were overwhelmingly due to airborne-droplet transmitted infections, with only seven due to mostly faecal-oral transmitted infections [10]. Accordingly, we assumed that on an annualised basis 𝐷_𝑢,𝑎,base_ would be split into 30% due to COVID-19 (see above), with the remaining 70% divided between other endemic airborne-droplet (95% of 70% = 66%) and faecal-oral (5% of 70% = 4%) diseases. We further allocated the above proportions to specific diseases and age groups based on observational or disease burden modelling studies of Gaza, where available, the West Bank alternatively, or, as a last resort, the Middle East region (Table S4). Lastly, we reviewed MoH data or, failing this, observational studies from Gaza, the West Bank or the Middle East to attribute annual caseload (and thus mortality) into monthly fractions and thus compute 𝜔_𝑢,𝑡_ (Table S5).

#### Transmissibility, case-fatality and epidemic probability

We combined structured expert elicitation (SEE) [48] with values from the literature to quantify the basic reproduction number ℛ_0,𝑢_ and the age-specific CFR 𝜇_𝑢,𝑎_, which in turn are needed to construct 𝜑_𝜆,𝑢,𝑡_ and 𝜑_𝜇,𝑢,𝑡_; and the probability of an epidemic occurring 𝑝_𝑢_. Seven infectious disease and/or ‘humanitarian’ epidemiology experts were invited via email to answer an online structured questionnaire on the ODK platform after consenting to participate in the study. They were asked for their estimates of the probability of an epidemic occurring during the next six months for each epidemic-prone pathogen included in the analysis; the ℛ_0_ of measles and cholera; and the CFR among children aged 12 to 59mo of symptomatic measles and cholera cases. Measles and cholera were selected as exemplars of airborne-droplet and faecal-oral transmitted infections, respectively, since these share key commonalities including risk factors and (mostly) treatment requirements (antibiotics and respiratory support for the former; antibiotics and rehydration for the latter). This was done to avoid an unmanageably long questionnaire and pre-empt repetitive or uninformative answers. The panel was provided with information on the three scenarios (Tables S2, S3), including susceptibility to epidemic-prone infections as estimated above, estimated values for epidemiological risk factors, the occurrence and timing of epidemics in past crises (Table S9); and the published ranges of ℛ_0_ and 𝜇 for measles and cholera (Tables S10, S11).

For each quantity and scenario, experts were asked to provide estimates for three probability quintiles: ‘lowest-reasonable’ (10th percentile, or ‘there is a 10% chance that the value will be even lower’), ‘most-likely’ (50^th^ percentile) and ‘highest-reasonable’ (90% percentile, or ‘there is <10% chance that the value will be even higher’). Experts were also asked to answer 10 ‘calibration’ questions in order to assess expert reliability, using the same 10-50-90% scheme: these questions concerned key infection parameters and have a strong evidence-based answer. The answers to the calibration questions were used to generate calibration and information scores for each expert, as shown by Cooke et al. [48]. The two scores were multiplied together to generate a weight for each expert, and compute weighted mean probability distributions (Figures S5-S7).

For measles we found that experts provided ℛ_0_ values implausibly similar to or lower than plausible estimates pre-war, based on European and North American cities (12 to 18). After consultation with a separate group of infectious disease experts, we decided to override the elicitation panel and revise the distribution of ℛ_0, measles_ as Runif(16,20), Runif(20,24) and Runif(24,28) for the ceasefire, status quo and escalation scenarios during months 1-3, and Runif(18,22), Runif(22,26) and Runif(26,30) respectively during months 4-6.

During model implementation, we sampled from the empirical cumulative probability distributions (ECDF) of ℛ_0_ and 𝜇 of measles and cholera. For other epidemic-prone infections, we computed ℛ_0,𝑢_ based on the relative position along its published range that corresponded to the relative position of their exemplar disease’s random ℛ_0_ value. For example, for diphtheria

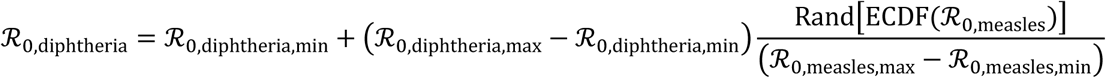

For endemic infections, we pre-defined a plausible range within which the pre-war (subscript _base_) theoretical transmissibility of both exemplar diseases would lie. For measles, we took the published range of ℛ_0_ in European and North American cities (i.e. ℛ_0,measles,base_ = 12 to 18) [49], while for cholera we reasoned that baseline transmissibility would have been just below the minimum values estimated during actual cholera outbreaks (Table S10; ℛ_0,cholera,base_ = 0.8 to 1.0); this reflects the observation that in the Middle East, cholera has only taken hold in crisis-affected settings and not where public health services were functional [50–53]. We then computed each simulation run’s 𝜑_𝜆,𝑢,𝑡_ as 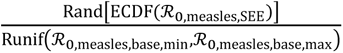 for all airborne-droplet transmitted infections, and 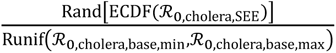 for faecal-oral transmitted infections.

We applied the same scaling to draw random values of CFR and 𝜑_𝜇,𝑢,𝑡_. Since the expert-elicited CFR referred to the age group 12-59mo, we computed CFR for other age groups based on published estimates of their relative difference from this reference age group (Table S11).

### Ethics

The project received approval from the Johns Hopkins Bloomberg School of Public Health Institutional Review Board and the London School of Hygiene & Tropical Medicine Research Ethics Committee (ref. 29926).

## Results

### Overall estimates

Under counterfactual no-crisis conditions, we estimated that 158 (95% confidence interval, CI 122 to 197) endemic infectious disease deaths would have occurred during the four-month retrospective analysis period (7 October 2023 to 6 February 2024), which comprised the seasonal peak in acute respiratory infections, and 191 (95%CI 156 to 237) during the six-month projection period (7 February to 6 August 2024). We assumed that no epidemic deaths would have occurred.

We estimated that 2056 (95%CI 517 to 5743) endemic infection deaths would occur during the retrospective period, i.e. 13.0 (95%CI 4.2 to 29.2) times higher than the counterfactual; corresponding levels over the projection period were 960 (95%CI 146 to 3529), 1301 (95%CI 195 to 3975) and 1538 (95%CI 236 to 4478) under the ceasefire, status quo and escalation scenarios, respectively, or 5.0, 6.8 and 8.1 times higher than baseline considering the point estimates. Resulting excess mortality is summarised in Table 3. Should epidemics occur during the projection period, we projected that these could cause between 5026 and 11,457 excess deaths depending on the scenario, but with very wide CIs extending to nearly 130,000 under the reasonable-worst scenario.

**Table 3.** Estimated numbers of *excess* deaths due to endemic and epidemic infectious diseases, by period and scenario. Values are the mean estimate and the 95% uncertainty interval.

| Cause | Retrospective period†<br>(7 Oct 2023 to 6 Feb 2024) | Projection period<br>(7 Feb to 6 Aug 2024) |  |  | Total analysis period<br>(7 Oct 2023 to 6 Aug 2023) |  |  |
| --- | --- | --- | --- | --- | --- | --- | --- |
|  |  | ceasefire | status quo | escalation | ceasefire | status quo | escalation |
| Endemic infectious diseases | 1832<br>(138 to 5442) | 1,522<br>(-74 to 5,746) | 2,118<br>(-29 to 7,522) | 2,721<br>(9 to 9,967) | 3354<br>(64 to 11,188) | 3950<br>(109 to 12,964) | 4553<br>(147 to 15,409) |
| Epidemic-prone infectious diseases | 0<br>(0 to 0) | 5,026<br>(0 to 58,741) | 8,463<br>(0 to 100,323) | 11,457<br>(0 to 128,917) | 5,026<br>(0 to 58,741) | 8,463<br>(0 to 100,323) | 11,457<br>(0 to 128,917) |

† Parameterised as per the status quo scenario.

### Endemic diseases

COVID-19, influenza and pneumococcal disease were the leading expected causes of death (Table 4), but with a lower toll in the projection period due to these diseases’ seasonality pattern.

**Table 4.** Projected excess deaths (95% confidence interval) due to endemic infections, by disease, scenario and period.

| Disease | Retrospective period<br>(7 Oct 2023 to 6 Feb 2024) | Projection period (7 Feb to 6 Aug 2024) |  |  |
| --- | --- | --- | --- | --- |
|  |  | ceasefire | status quo | escalation |
| bacterial gastroenteritis | 45 (6 to 105) | 38 (4 to 120) | 64 (5 to 165) | 75 (7 to 179) |
| COVID-19 | 342 (23 to 1024) | 582 (-34 to 2,219) | 797 (-17 to 2,894) | 1,029 (-3 to 3,866) |
| *Haemophilus influenzae* type b disease | 0 (0 to 0) | 0 (0 to 0) | 0 (0 to 0) | 0 (0 to 0) |
| influenza, para-influenza | 687 (49 to 2069) | 252 (-15 to 956) | 346 (-8 to 1,268) | 431 (-3 to 1,616) |
| viral gastroenteritis (other than rotavirus) | 45 (6 to 105) | 38 (4 to 120) | 64 (5 to 165) | 75 (7 to 179) |
| pneumococcal disease | 479 (35 to 1448) | 486 (-28 to 1,864) | 668 (-14 to 2,408) | 889 (0 to 3,347) |
| rotavirus | 23 (3 to 52) | 19 (2 to 60) | 32 (3 to 82) | 37 (3 to 89) |
| RSV | 212 (15 to 638) | 107 (-6 to 407) | 148 (-3 to 540) | 185 (-1 to 692) |
| **total** | **1832 (138 to 5442)** | **1,522 (-74 to 5,746)** | **2,118 (-29 to 7,522)** | **2,721 (9 to 9,967)** |

We estimated that during the retrospective period 909 (95%CI 187 to 2687) excess deaths occurred among persons ≥60yo (47.9%) and 288 (95%CI 62 to 792) among children ≤59mo (15.2%). Under the ceasefire scenario, 913 (95%CI −50 to 3,474) excess deaths were projected among persons ≥60yo (60.0%), and 197 (95%CI −2 to 714) among children ≤59mo (12.9%). The corresponding figures for the status quo and escalation scenarios were 1256 (95%CI −23 to 4520) and 1,627 (95%CI 0 to 6061) for persons ≥60yo (59.3% and 59.8%), and 289 (95%CI 4 to 950) and 361 (95%CI 9 to 1203) among children ≤59mo (13.6% and 13.3%).

### Epidemic-prone diseases

Table 5 shows projected deaths due to different epidemic-prone infections under different scenarios (as the assumed counterfactual was zero, all mortality is excess). Cholera, measles, polio (both wild-type and vaccine-derived) and meningococcal meningitis accounted for the greatest mortality threat. Measles was expected to cause moderate-size epidemics at most due to relatively high baseline herd immunity, while immunity appeared low for vaccine-type polio (Figure S4).

**Table 5.** Projected mortality due to epidemic-prone infections during the projection period, by disease and scenario. Values are the mean estimate and the 95% uncertainty interval.

| Disease | Scenario |  |  |
| --- | --- | --- | --- |
|  | ceasefire | status quo | escalation |
| bacterial dysentery (*S. dysenteriae* type 1) | 0 (0 to 3) | 0 (0 to 6) | 0 (0 to 7) |
| cholera | 3,595 (0 to 50,550) | 6,299 (0 to 91,671) | 8,071 (0 to 119,661) |
| diphtheria | 0 (0 to 3) | 0 (0 to 5) | 1 (0 to 11) |
| hepatitis A | 0 (0 to 0) | 0 (0 to 0) | 0 (0 to 0) |
| hepatitis E | 2 (0 to 27) | 2 (0 to 38) | 3 (0 to 48) |
| measles | 260 (0 to 3,211) | 454 (0 to 5,192) | 793 (0 to 9,662) |
| meningococcal meningitis | 24 (0 to 265) | 48 (0 to 570) | 143 (0 to 1,762) |
| pertussis | 0 (0 to 0) | 0 (0 to 0) | 0 (0 to 1) |
| polio - vaccine-derived | 639 (0 to 8,007) | 934 (0 to 12,438) | 1,394 (0 to 20,909) |
| polio - wildtype | 506 (0 to 6,187) | 724 (0 to 10,050) | 1,050 (0 to 14,906) |
| typhoid fever | 0 (0 to 4) | 0 (0 to 5) | 0 (0 to 6) |
| **total** | **5,026 (0 to 58,741)** | **8,463 (0 to 100,323)** | **11,457 (0 to 128,917)** |

Among persons ≥60yo, 527 (95%CI 0 to 6340; 10.5%), 884 (95%CI 0 to 10,942; 10.5%) and 1176 (95%CI 0 to 14,086; 10.3%) deaths were projected to occur in the ceasefire, status quo and escalation scenarios, respectively. Corresponding projections for children ≤59mo were 571 (95%CI 0 to 6846; 11.3%), 1023 (95%CI 0 to 12,283; 12.0%) and 1451 (95%CI 0 to 16,187; 12.3%). A relatively larger share of deaths was projected among adults, mainly reflecting the high CFR of cholera in this age group.

Epidemic projections carried high uncertainty: model output distributions were heavily left-skewed towards 0 deaths (the most common result given that 𝑝_𝑢_ < 0.50 for all diseases). We therefore also present the probability of epidemics due to different pathogens, or all pathogens combined, reaching thresholds of 100, 1,000 or 10,000 deaths (Figure 3). The probability of epidemics causing ≥1,000 deaths rose from 23% to 27% and 34% under the ceasefire, status quo, and escalation scenarios, respectively.

**Figure 3.**
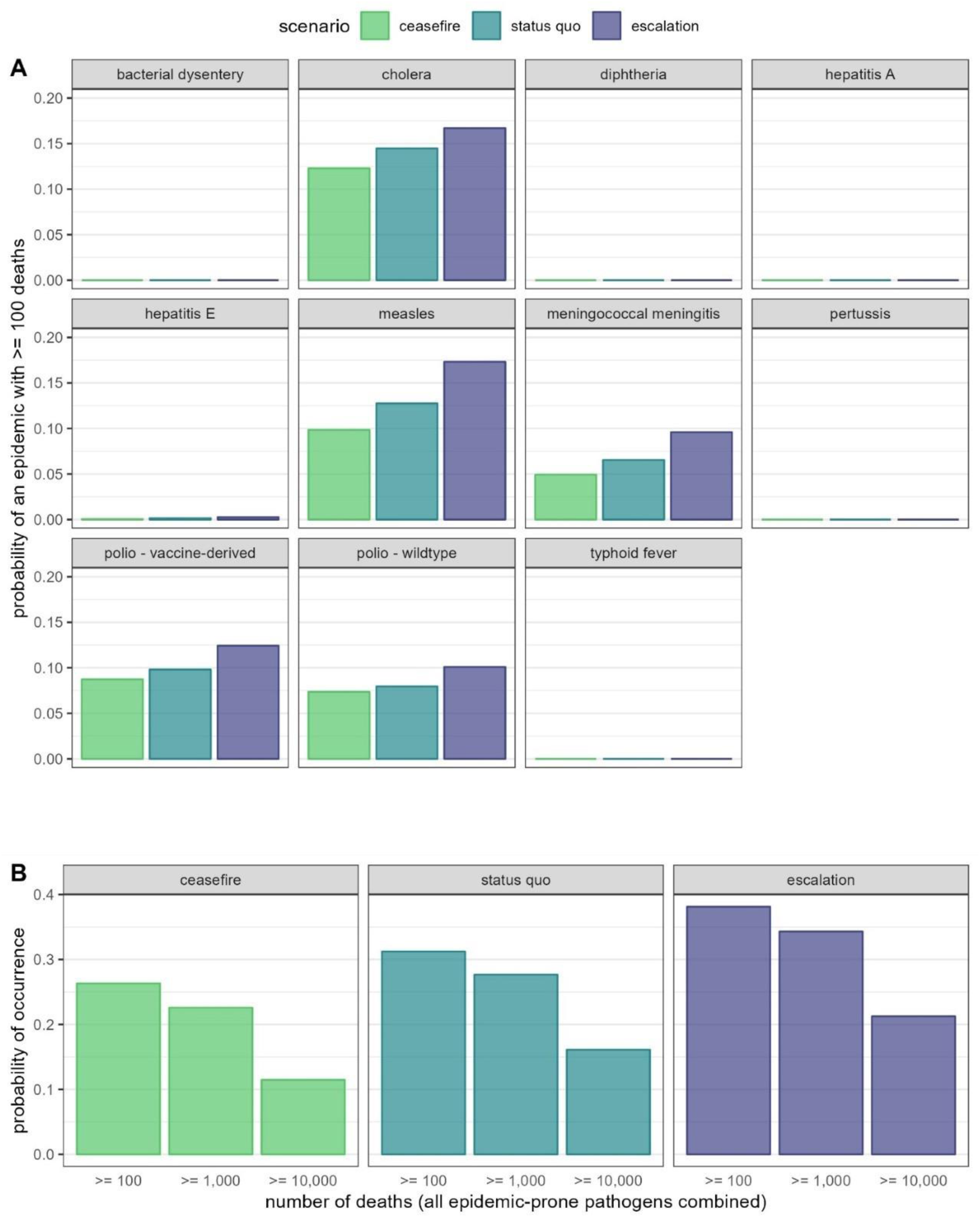
<u>Panel A</u>: Estimated probability of occurrence of an epidemic featuring ≥ 100 deaths, by disease and scenario, over the six-month projection period. <u>Panel B</u>: Estimated probability of exceeding specific death toll thresholds due to all epidemic-prone pathogens combined, by scenario, over the six-month projection period.

## Discussion

To our knowledge this is the first study attempting comprehensive, age-stratified projection of infectious disease mortality across multiple pathogens in a real-time humanitarian response. We notably accounted for both transmissibility and case-fatality increases, seasonality and the long-term dynamics of population susceptibility. Our approach is distinct from other models of future mortality that specifically feature infectious diseases. The Lives Saved Tool (LiST) [54] models mortality deterministically among children under 5yo and pregnant women as a function of the coverage of various interventions (e.g. clean water), but does not readily accommodate crisis-emergent risk factors such as overcrowding; diarrhoea, HIV, pneumonia and malaria are modelled specifically, but epidemics aren’t. The Global Burden of Disease (GBD) project [55] retrospectively estimates and forecasts deaths and disability due to a wide range of infections including epidemics, with specific methods for each, largely based on statistical and geospatial models: its analysis horizon is typically multi-year and, to our knowledge, has not been adapted to consider the short timeframe and risk factors specific of crises. Both LiST and GBD, however, explicitly incorporate intervention coverage and effectiveness, which, with the exception of susceptibility analysis, we merely featured as factors for SEE to consider.

It is unclear how well our estimates performed in comparison to what actually happened in Gaza. While the retrospective estimates (up to 6 February 2024) could fairly be benchmarked against empirical evidence, no ground-based study of non-injury mortality is available. High diarrhoea caseloads were reported early in the crisis [56] and an increase in bloody diarrhoea consultations was apparent [57], but in a relatively well-nourished population (at least during the first 3-4 months of the war) these could have featured a low CFR; our estimates suggest that respiratory infections would have had a much greater impact during this period (overlapping the cold season). Scenario projections should not be judged on their predictive accuracy, as they depend on scenario choices that may not materialise. Nevertheless, it is notable that Gaza had not experienced major epidemics as of April 2025, with the only confirmed outbreak due to vaccine-derived type 2 poliovirus in July 2024 (3 confirmed symptomatic cases) and followed by mass vaccination campaigns. Our analysis did suggest a particularly low level of immunity to vaccine-derived polio infection (Figure S3), reflecting the withdrawal of type 2 from recent vaccination schedules. We speculate that other epidemics (cholera, measles) did not occur due to a combination of chance (both would have had to be imported), moderate levels of malnutrition and high herd immunity (for measles). Surveillance data also suggest an exceptional increase in acute jaundice during early 2024 [57], consistent with an epidemic, but with an unclear aetiologic agent.

## Limitations

Our models relied on a large number of disease-related parameters that all carry some uncertainty, especially for diseases and vaccines that have been comparatively under-researched. We did not have time and resources to conduct systematic reviews of all these parameters, though we relied on reviews where available. While it is unlikely that resulting error would have caused large bias overall (i.e. it is implausible that we would have over-or under-estimated parameter values across all diseases), we have not fully represented this uncertainty in our estimates, especially for parameters assumed to not vary by age. Similarly, susceptibility analysis is reliant on robust assumptions about vaccine effectiveness and prior force of infection, which, taken together, determine the final size of epidemics. The latter was difficult to estimate, though it is unlikely, based on sensitivity analysis, to have biassed our findings other than for hepatitis A, which we incorrectly treated as epidemic-prone, not accounting for its high 𝜆 in Gaza, even pre-war [58]. Future implementations of the method could rely on infection-specific dynamic or catalytic models tuned to similar settings where more data are available to quickly provide a more informed estimate of immunity status even if local 𝜆 is unknown.

For some older-generation vaccines (e.g. polio), effectiveness against infection or disease is not particularly well documented; moreover, we held effectiveness fixed rather than varying it to allow for uncertainty in published reports. SEE produced implausible estimates of transmissibility for one of the two exemplar diseases, suggesting problems with the presentation of the questions or their understanding. In future iterations of the method, experts may need to receive more explicit and clear instructions or may need more support in the form of pre-briefings to fully comprehend the information and what is being asked of them [59]; calibration questions may need to also be more discriminant.

Beyond the accuracy of data inputs, the SEIR epidemic models do not account for (i) spatial heterogeneity (for example, it is plausible that there were pockets of particularly high susceptibility or transmissibility in the worst-off communities in Gaza); and (ii) interventions that may be implemented to control epidemics, e.g. reactive vaccination (we did however incorporate availability of curative care in our scenarios). Moreover, the critical feedback loop of malnutrition and infection was incompletely featured in our analysis, relying solely on elicitation experts’ incorporating projected malnutrition and breastfeeding disruptions into their judgment, alongside many other risk factors: it is unclear to what extent experts were able to do so.

## Conclusions

Projecting the mortality risk of specific infectious diseases could enhance situational awareness and planning in humanitarian response: indeed, recent foreign aid cuts warrant ever-more efficient and targeted use of shrinking resources. The initial experience reported here provides a foundation for a more systematic and crisis-generic scenario-based projection approach. However, several improvements to the models are warranted, among which the most critical are mechanistic treatment of the malnutrition-infection loop; better methods for expert elicitation; and more consistent propagation of uncertainty. Working with end users on optimal ways to convey the complex statistical estimates (e.g. concepts of epidemic probability) may also be beneficial. In most crises, the rich baseline data available for Gaza would likely be missing, necessitating more flexible data inputs, e.g. burden-of-disease models where ground data are unavailable. Lastly, the method required extensive local expertise, mathematical modelling inputs, software coding and evidence reviews, potentially limiting its applicability where these resources are not available within the short timeframe required for humanitarian decision-making. It is likely that its systematic application across crises would require sustained investments in standing expertise.

## Statements

### Funding

This research was conducted with the support of the UK Humanitarian Innovation Hub (UKHIH) and its donor, the UK Foreign, Commonwealth & Development Office (FCDO).

## Data Availability

All source data and analysis code required to replicate the analysis are publicly available on https://github.com/Gaza-projections/gaza_projections/tree/main/gaza_infections.

## Acknowledgments

The Gaza Health Impact Projections Working Group includes the following co-authors of the overall report and specific modules: Sarah Aly, Shatha Elnakib, Hannah Tong, Tak Igusa and Oona MR Campbell. It also includes colleagues and students who volunteered time to collect data for this analysis: Lina Abdulsamad, Berthe Abi Zeid, Kirsty Andresen, Rachel Arrundale, Sarah Arunachalam, Aamena Valiji Bharmal, Marleen Bokern, Aparna Sri Dasaraju, Rebecca Debruyn, Sarah Dickson, Alexander Frias, Anika Gnaedinger, Tarek Jaber, Dana Jamaluddine, Laukhika Kasetty, Amelie Niemann, Nadeem Obaydou, Lisa Maria Persson, Maria Jose Sanchez Alva, Aline Semaan, Rezwana Uddin, Francesco Venuti, Zeenat Williams, Sarah Zaidi.

We gratefully acknowledge project management support by Mortala Ndow and Elmy Thompson, in-kind contributions towards questionnaire design, web development and graphics layout by Chrissy H. Roberts, Fatima Ali Ahmed, Tatiana Elghossain, Ghaleb Hamadi, Ruba Saleh and Teddy Zeenny and contributions to software code by Emilie Finch, Sebastian Funk and Pratik Gupte.

The following experts provided advice on infectious disease estimation methods and/or information inputs and are gratefully acknowledged: John Edmunds, Paul Fine, Adam Kucharski, Punam Mangtani, Carmen Tamayo Cuartero, Laura Rodrigues, and 3 more experts who preferred to remain anonymous.

Experts providing elicitation estimates were Laith Abu-Raddad, Bhargavi Rao, and five more experts who preferred to remain anonymous.

Lastly, we are thankful to the following agencies for facilitating access to data used in the analysis: United Nations Children’s Fund, United Nations Office for the Coordination of Humanitarian Affairs, United Nations Population Fund, United Nations Relief and Works Agency for Palestine Refugees in the Near East, World Health Organization.

## Notes

### Competing Interest Statement

The authors have declared no competing interest.

### Author Declarations

All data used for the study were publicly available and previously collected, and are curated at https://github.com/Gaza-projections/gaza_data_sources. See citations inside the manuscript.

