## Supplementary Information for "Projecting excess mortality due to infectious diseases during a crisis: methods and application to the Gaza Strip, 2023-2024"

### List of tables and figures

[Table S1. Gaza population by age and sex, based on UNFPA 2023 projections [1].	3](#_Toc200554437)

[Figure S2. Number of daily contacts between age groups, based on van Zandvoort et al [12].	8](#_Toc200554449)

[Figure S3: **A** Vaccine coverage used in the immunity model for epidemic-prone diseases, from 2000-2025. Values taken from MoH Gaza Report 2022 and UNICEF Palestinian Multiple Indicator Cluster Surverys (MICSs) 2010, 2014, and 2018. Where there are two data sources, these are averaged. Missing values are imputed by assuming values coverage from the latest data source. These back propagated values at 2000 give a similar result to coverage reported in MIC 2000 (which would cover the model year of 1999-2000). **B** Depicts the vaccine schedules used in immunity model, to simplify the computation doses between 2 months and 6 months are treated as a single vaccination at 3-4 months.	13](#_Toc200554450)

### Study scope

#### Population denominators

To obtain sex- and age-specific population denominators (Table S1) we used United Nations Population Fund (UNFPA) May 2023 estimates for Gaza, with some modifications, explained below. The UNFPA estimates are based on the Palestine census held on 1 December 2017, and provide five-year age groups for males and females separately for the age groups of 0-4 to 75-79, and 80+ [1]. The total population matches with the Palestine Central Bureau of Statistics (PCBS) projection for 2023, although the latter does not disaggregate by sex- or age-groups [2].

To obtain the age distribution by year for children under five, we used the relative proportions contained in the 2022 PCBS estimates, which were given in single years for those 0-4yo. To further break down the infant group into neonates (<1mo) and 1-11, we assumed an equal proportion by month (i.e. 1/12 and 11/12).

**Table S1.** Gaza population by age and sex, based on UNFPA 2023 projections [1].

| **Age†** | **Population** | | |
| --- | --- | --- | --- |
|  | **male** | **female** | **total** |
| <1mo | 2,959 | 2,851 | 5,810 |
| 1-11mo | 32,545 | 31,366 | 63,911 |
| 12-59mo | 136,078 | 131,258 | 267,336 |
| 5-9yo | 145,276 | 139,182 | 284,458 |
| 10-14yo | 141,660 | 135,532 | 277,192 |
| 15-19yo | 120,553 | 115,384 | 235,937 |
| 20-29yo | 195,265 | 188,511 | 383,776 |
| 30-39yo | 151,718 | 150,342 | 302,060 |
| 40-49yo | 90,426 | 91,191 | 181,617 |
| 50-59yo | 60,013 | 58,011 | 118,024 |
| 60-69yo | 33,911 | 33,943 | 67,854 |
| 70-79yo | 14,216 | 15,890 | 30,106 |
| 80-100yo | 3,293 | 5,170 | 8,463 |
| **Total** | **1,127,913** | **1,098,631** | **2,226,544** |

† (mo) month old (yo) year old.

#### Projection scenarios

**Table S2.** Qualitative description of the three scenarios.

| Scenario | Intensity and typology of military activity and population movement | Occurrence and duration of pauses/ceasefires | Humanitarian space and operational adaptation |
| --- | --- | --- | --- |
| Ceasefire | - A permanent ceasefire occurs, and ariel bombing stops, and violence reduces. - Gaza remains under blockade with border checks and military policing at the border. - Telecommunication resume - The population starts to return home, but the majority remain in shelters/informal settlements due to destroyed dwellings. | - Ceasefire is agreed to over the entire period | - A large influx of humanitarian assistance enters Gaza but is limited at the beginning & increases over time as logistics improve in Gaza (roads, storage); - More goods allowed in. - The international community supports an emergency response. Rebuilding and repairing damaged health facilities, homes and infrastructure. - Health services at all levels can operate safely with adequate access, supplies and quality improving over time; primary health services expand to areas where no services exist; paucity of specialized healthcare (e.g. surgeries) and functioning referral pathways occur within and outside of Gaza over time. - Large vaccination campaigns and other distributions occur. - Water, sanitation and hygiene (WASH) improve in shelters and open areas, reducing the probability of epidemics. - Food provision increases. - Humanitarian actors adapt their operations by shifting activities closer to the populations and with international community works to repair damaged healthcare services and hospitals to increase access and quality of services; this will be slow due to extensive damage. - International communities work to rebuild housing infrastructure to eventually reduce overcrowding; this will also be slow to occur due to extensive damage. |
| Status quo | - Aerial bombing continues, but the military campaign evolves towards more urban ground warfare. - An offensive against South Gaza occurs. - Humanitarian pauses reduce violence, occasional bombings and ground attacks occur, then continue as soon as pauses end. - Gaza remains under *de facto* military control with stringent border checks and limited goods entry. - A near-total telecommunications blackout across the Gaza Strip most of the days. - Displacement persists and as people continue to move south. - People shelter in open areas (streets,) as shelters are full; most are unable to return to their houses (many of which are destroyed). | - Two or three humanitarian pauses are agreed, each of about 5-7 days. | - A limited increase in aid being allowed to enter Gaza, in particular food, but all items remain insufficient (water, fuel, medicines and supplies). - Humanitarian action remains very constrained by operating restrictions, and large pockets of Gaza’s Palestinian population are only intermittently accessible if at all, particularly in North and Middle areas. - Functionality of health services remains at current low levels including low or unavailable stocks of essential medication and supplies; referrals within or outside of Gaza are extremely limited. Quality of services, and intervention coverage, remain very constrained. - Epidemics and acute malnutrition are likely to occur. - Some adaptation of humanitarian services in South, e.g. by moving health services closer to Internally displaced persons (IDPs), optimizing supply chains, implementing vaccination campaigns and making health facilities more resilient to attacks. |
| Escalation | - Military campaign intensifies during the period including aerial bombardment and ground force operations with increased intensity shifting the focus to the crowded area in the South of Gaza and resume in the North. - Gaza remains under *de facto* military control with stringent border checks and limited goods entry. - A total telecommunications blackout across the Gaza Strip. - Continued large-scale displacement, now into more open areas as shelters are full (increasing displacement requiring more outdoor tents). - More health facilities become partially -or non-functional and many people are in areas where there were fewer health facilities to begin with. - Most of the population will have less access to care due to limited hospitals, clinics, and transportation to healthcare facilities. | - None | - The humanitarian and health situation worsens in all areas of Gaza. - Humanitarian space constrained – worsened insecurity for humanitarian actors and Gazan health workers; limited services provided compared to current situation; humanitarian community does not/cannot respond to these challenges by adapting. - North has with very limited assistance provided. - Closed or reduced health facilities at all levels with stockouts of essential medicines and equipment; difficulties in providing health care to those in open/remote places. Clear deterioration of health services re: access, supply, quality; epidemics likely to occur. - The amount of water, food, fuel, and medicines is even more limited. |

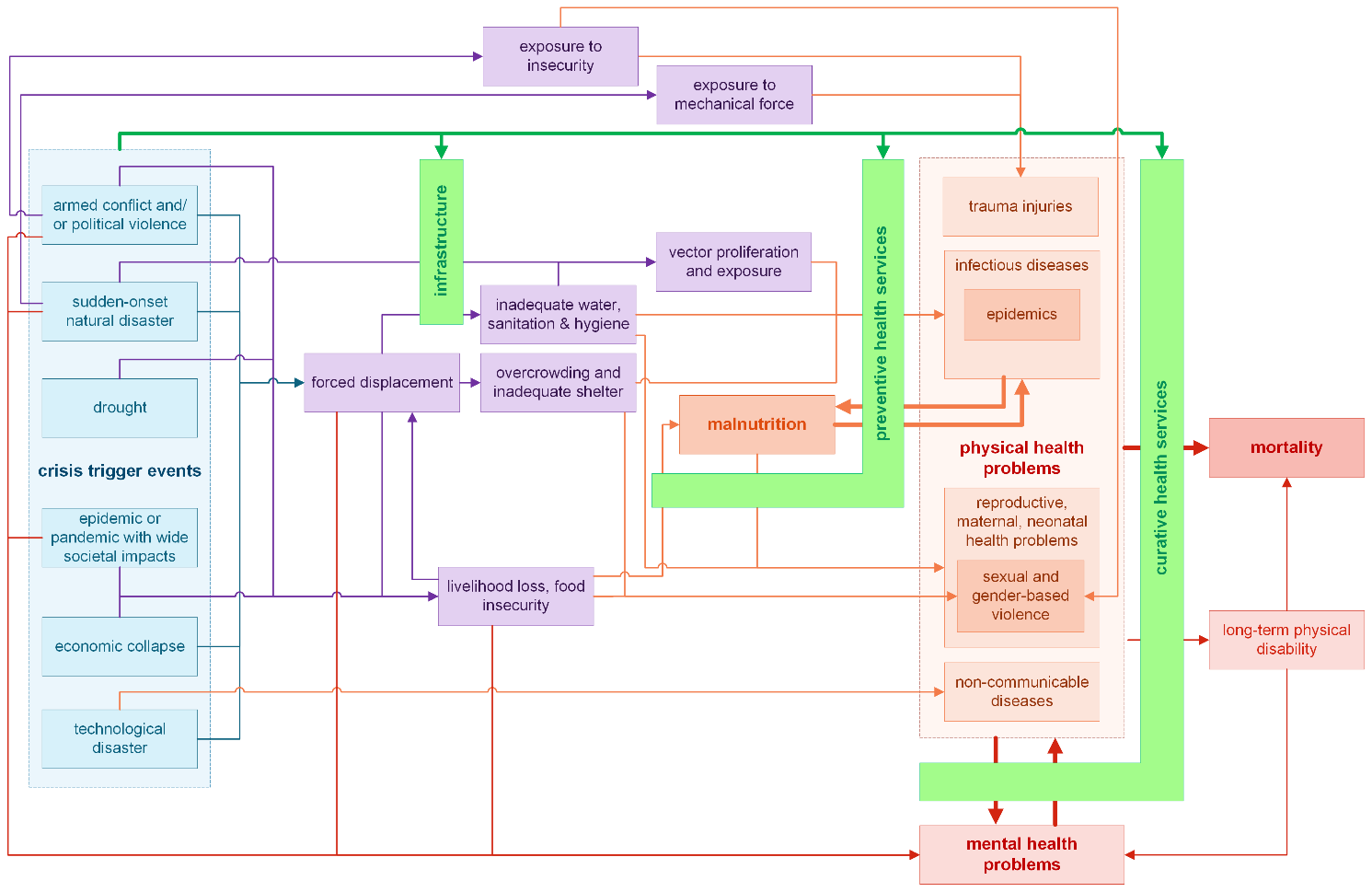

**Figure S1.** Proposed causal framework of factors leading to (excess) morbidity, disability and mortality in a crisis. Infectious diseases are outlined in dark red.

**Table S3.** Assumed value of key parameters relevant to the transmissibility and/or case-fatality of infectious diseases, by scenario.

| **Parameter** | **Value by scenario** | | | **Notes** |
| --- | --- | --- | --- | --- |
|  | ceasefire | status quo | escalation |  |
| Mean litres of water available per person per day (L/p/d) | 20-40 | 6-10 | 3-6 | 8.8 L/p/d in shelters [3] against Sphere standard of survival 7.5 and minimum of 15 [4]. Access difficult and North has very poor access. No comprehensive water quality testing has been conducted, there are concerns over water quality in Gaza strip [5]. |
| Number of persons per toilet (p/toilet) | 130-350 | 450-800 | 580-1000 | 486 people per toilet [6] while the standard during the first phases of a rapid-onset crisis is with a minimum ratio of 1 communal toilet per 50 people [4] Some toilets may not be functional. Reporting of open defecation [7]. |
| Percentage of people with access to soap | 50-70% | 30-40% | 10-30% | 142,669 hygiene kits (including soap) have been distributed since October (~35% of target) [5]. No other standard indicators currently reported for handwashing with soap. |
| Proportion of recommended daily intake met through food aid as compared to 2213 kcal/day | 1.10-1.30 | 1.00-1.20 | 0.80-1.00 | See nutrition module in wider projections project’s report 1, methods annex: <https://gaza-projections.org/docs/gaza_projections_methods_annex.pdf> |
| Prevalence of moderate or severe acute malnutrition as per WHO definitions among children aged 6 to 59 months | 8-10% | 13-16% | 20-45% |  |
| Percentage of infants who are exclusively breastfed: |  |  |  |  |
| aged below 1 month | 30-40% | 30-40% | 20-25% |  |
| aged below 6 months | 25-30% | 25-30% | 15-20% |  |
| Percentage of people living in communal shelters or tented camps | 50-60% | 70-80% | 90-95% | UNRWA schools and governmental shelters 1.9 million [8]. |
| Mean number of people per communal shelter or tented camp | 4500-5000 | 7500-8000 | 8500 -10,000 |  |
| Percentage of people living in inadequately winterized/ heated shelters/houses | 55-60% | 90-95% | 90-95% |  |
| Percentage of infants who received their third dose of routine vaccination | 30%-60% | 10-20% | 5-10% | Based on access to hospitals/ PHC [9], supply and quality of services provided [10]. |
| Percentage of the population with access to outpatient public curative health facilities that are functional (for antimicrobials or oral rehydration salts) | 50-70% | 20-40% | 5-10% |  |
| Percentage of the population with access to inpatient public departments that are functional and able to administer rehydration | 20-70% | 10-15% | 5-10% |  |
| Percentage of the population with access to inpatient public departments that are functional and able to administer antimicrobials | 20-70% | 10-15% | 5-10% |  |
| Percentage of the population with access to inpatient public departments that are functional and able to administer respiratory support including oxygen | 20-70% | 5-10% | 1-5% | Based on access to hospitals [9], supply and quality of services provided. Very limited supply of oxygen [11]. |

### Analytic framework

#### Epidemic-prone infections model

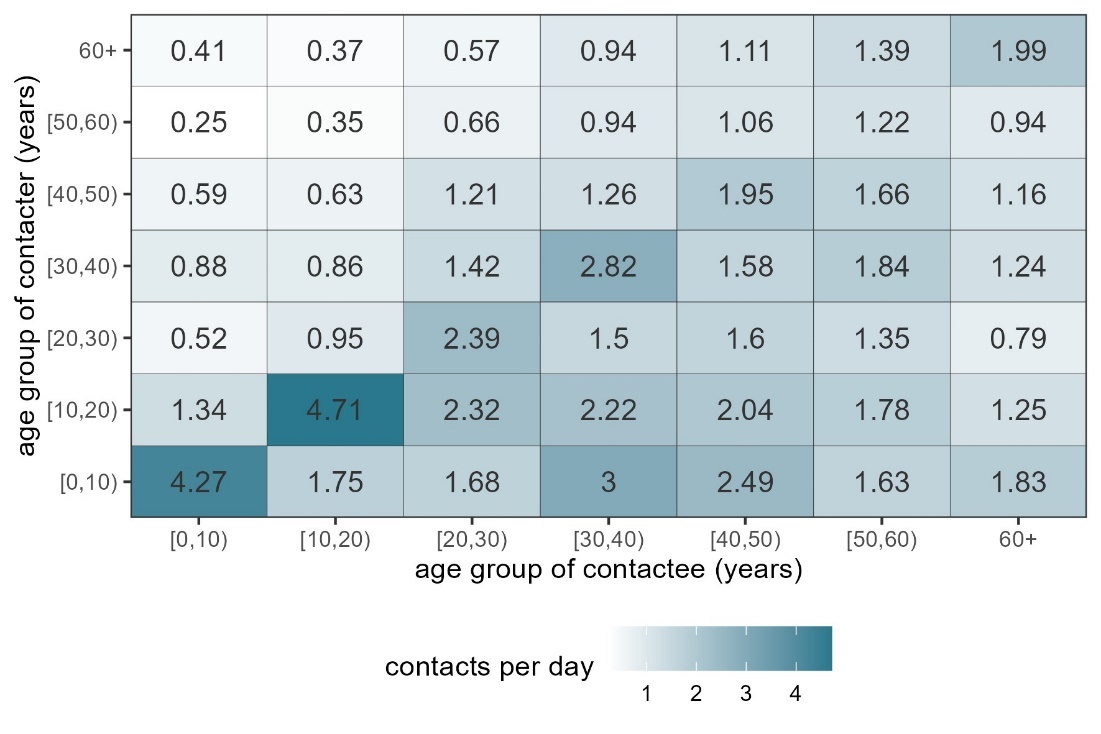

**Figure S2.** Number of daily contacts between age groups, based on van Zandvoort et al [12].

#### Counterfactual (baseline) mortality by age, sex and season

**Table S4.** Assumed disease-specific and age distribution of baseline endemic infectious disease mortality.

| **Infectious disease** | **Proportion of deaths** | **Source** | **Age distribution of mortality** | | | | | | | | | | | | | **Source** |
| --- | --- | --- | --- | --- | --- | --- | --- | --- | --- | --- | --- | --- | --- | --- | --- | --- |
|  |  |  | <1  mo | 1-11  mo | 12-59 mo | 5-9  yo | 10-14  yo | 15-19  yo | 20-29  yo | 30-39  yo | 40-49  yo | 50-59  yo | 60-69  yo | 70-79  yo | ≥80  yo |  |
| **Airborne-droplet other than COVID-19 (66%)** | | | | | | | | | | | | | | | | |
| Hib disease | 0.00 | [13] | 0.100 | 0.600 | 0.300 | 0.000 | 0.000 | 0.000 | 0.000 | 0.000 | 0.000 | 0.000 | 0.000 | 0.000 | 0.000 | [14] |
| pneumococcal disease | 0.50 | [15] | 0.002 | 0.023 | 0.096 | 0.024 | 0.015 | 0.010 | 0.022 | 0.035 | 0.051 | 0.088 | 0.151 | 0.189 | 0.295 | [15] |
| RSV | 0.13 | [16] | 0.011 | 0.124 | 0.520 | 0.152 | 0.008 | 0.008 | 0.000 | 0.024 | 0.000 | 0.016 | 0.023 | 0.040 | 0.073 | [17] |
| influenza, para-influenza | 0.37 | all other deaths | 0.000 | 0.003 | 0.014 | 0.030 | 0.015 | 0.019 | 0.061 | 0.131 | 0.138 | 0.249 | 0.069 | 0.110 | 0.160 | [17] |
| **Airborne-droplet, COVID-19 (30%)** | | | | | | | | | | | | | | | | |
| COVID-19 | 0.30 | see text | 0.000 | 0.000 | 0.010 | 0.000 | 0.000 | 0.000 | 0.013 | 0.023 | 0.033 | 0.082 | 0.191 | 0.293 | 0.355 | [18, 19] |
| **Faecal-oral (4%)** | | | | | | | | | | | | | | | | |
| rotavirus | 0.20 | [20] | 0.010 | 0.113 | 0.473 | 0.030 | 0.029 | 0.025 | 0.040 | 0.031 | 0.019 | 0.012 | 0.007 | 0.165 | 0.046 | [21] |
| other viral gastroenteritis | 0.40 | [21] | 0.010 | 0.113 | 0.473 | 0.030 | 0.029 | 0.025 | 0.040 | 0.031 | 0.019 | 0.012 | 0.007 | 0.165 | 0.046 | [21] |
| bacterial gastroenteritis | 0.40 | [21] | 0.100 | 0.600 | 0.300 | 0.000 | 0.000 | 0.000 | 0.000 | 0.000 | 0.000 | 0.000 | 0.000 | 0.000 | 0.000 | [21] |

**Table S5.** Assumed proportion of annual cases occurring within a given calendar month.

| **Infectious disease** | **Proportion of annual cases occurring within a given month** | | | | | | | | | | | | | **Source** |
| --- | --- | --- | --- | --- | --- | --- | --- | --- | --- | --- | --- | --- | --- | --- |
|  | Jan | Feb | Mar | Apr | May | Jun | Jul | Aug | Sep | Oct | Nov | Dec |  | |
| **Airborne-droplet transmitted** | | | | | | | | | | | | | | |
| Hib disease | 0.083 | 0.083 | 0.083 | 0.083 | 0.083 | 0.083 | 0.083 | 0.083 | 0.083 | 0.083 | 0.083 | 0.083 | No evidence of seasonality [13] | |
| pneumococcal disease | 0.083 | 0.083 | 0.083 | 0.083 | 0.083 | 0.083 | 0.083 | 0.083 | 0.083 | 0.083 | 0.083 | 0.083 | No evidence of seasonality [17] | |
| RSV | 0.316 | 0.140 | 0.158 | 0.111 | 0.022 | 0.009 | 0.007 | 0.007 | 0.007 | 0.002 | 0.033 | 0.187 | [17] | |
| COVID-19† | 0.154 | 0.523 | 0.042 | 0.003 | 0.001 | 0.001 | 0.184 | 0.089 | 0.003 | 0.000 | 0.000 | 0.000 | [18] | |
| Influenza, para-influenza | 0.338 | 0.163 | 0.137 | 0.054 | 0.022 | 0.004 | 0.001 | 0.001 | 0.003 | 0.005 | 0.031 | 0.240 | [17] | |
| **Faecal-oral transmitted** | | | | | | | | | | | | | | |
| rotavirus | 0.083 | 0.083 | 0.083 | 0.083 | 0.083 | 0.083 | 0.083 | 0.083 | 0.083 | 0.083 | 0.083 | 0.083 | No evidence of seasonality [22] | |
| other viral gastroenteritis | 0.083 | 0.083 | 0.083 | 0.083 | 0.083 | 0.083 | 0.083 | 0.083 | 0.083 | 0.083 | 0.083 | 0.083 | No data: assumed no seasonality | |
| bacterial gastroenteritis | 0.083 | 0.083 | 0.083 | 0.083 | 0.083 | 0.083 | 0.083 | 0.083 | 0.083 | 0.083 | 0.083 | 0.083 | No data: assumed no seasonality | |

† Observed seasonal pattern may reflect transition between pandemic and endemic conditions and ongoing emergence of new variants.

### Model parameterisation

#### Susceptibility estimates

**Table S6.** Differential equations governing the flow of people into and out of different immunity compartments and age groups.

| **Compartment and age group** | **Equation†** |
| --- | --- |
| Susceptibles ($S_{a}$): | |
| neonates (0mo) | $\frac{dS_{a}}{dt}=b_{t}\left( \frac{S_{15-44yo}}{N_{15-44yo}} \right)+R_{a,t}w-S_{a,t}\left( \lambda_{a,t}+\alpha_{a}+d_{a} \right)$ |
| ≥ 7mo (when maternal immunity ends) | $\frac{dS_{a}}{dt}=\left( S_{a-1,t} + M_{a-1,t} \right)\times\alpha_{a-1}\rho_{u}+\left( R_{a,t}+V_{a,t} \right)w-S_{a,t}\left( \lambda_{a,t}+\alpha_{a}+d_{a} \right)$ |
| all other age groups | $\frac{dS_{a}}{dt}=S_{a-1,t}\alpha_{a-1}\rho_{u}+\left( R_{a,t}+V_{a,t} \right)w-S_{a,t}\left( \lambda_{a,t}+\alpha_{a}+d_{a} \right)$ |
| oldest age group | $\frac{dS_{a}}{dt}=S_{a-1,t}\alpha_{a-1}\rho_{u}+\left( R_{a,t}+V_{a,t} \right)w-S_{a,t}\left( \lambda_{a,t}+d_{a} \right)$ |
| Naturally immune ($R_{a}$): | |
| neonates | $\frac{dR_{a}}{dt}=\left( S_{a,t}+V_{\sigma,a,t} \right)\lambda_{a,t}-R_{a,t}\left( w+\alpha_{a}+d_{a} \right)$ |
| all other age groups | $\frac{dR_{a}}{dt}=R_{a-1,t}\alpha_{a-1}\rho_{u}+\left( S_{a,t}+V_{\sigma,a,t} \right)\lambda_{a,t}-R_{a,t}\left( w+\alpha_{a}+d_{a} \right)$ |
| oldest age group | $\frac{dR_{a}}{dt}=R_{a-1,t}\alpha_{a-1}\rho_{u}+\left( S_{a,t}+V_{\sigma,a,t} \right)\lambda_{a,t}-R_{a,t}\left( w+d_{a} \right)$ |
| Immune following vaccination ($V_{\lambda,a}$): | |
| 1mo | $\frac{dV_{\lambda,a}}{dt}=\left( S_{a-1,t}+M_{a-1,t}+R_{a-1,t} + V_{\sigma,a-1,t} \right)\rho_{\lambda}-V_{\lambda,a,t}\left( w+\alpha_{a}+d_{a} \right)$ |
| 2-6mo | $\frac{dV_{\lambda,a}}{dt}=V_{\lambda,a-1,t}\alpha_{a-1}+\left( S_{a-1,t}+M_{a-1,t}+R_{a-1,t} + V_{\sigma,a-1,t} \right)\rho_{\lambda}-V_{\lambda,a,t}\left( w+\alpha_{a}+d_{a} \right)$ |
| older ages | $\frac{dV_{\lambda,a}}{dt}=V_{\lambda,a-1,t}\alpha_{a-1}+\left( S_{a-1,t}+R_{a-1,t} + V_{\sigma,a-1,t} \right)\rho_{\lambda}-V_{\lambda,a,t}\left( w+\alpha_{a}+d_{a} \right)$ |
| oldest age group | $\frac{dV_{\lambda,a}}{dt}=V_{\lambda,a-1,t}\alpha_{a-1}+\left( S_{a-1,t}+R_{a-1,t} + V_{\sigma,a-1,t} \right)\rho_{\lambda}-V_{\lambda,a,t}\left( w+\alpha_{a} \right)$ |
| Immune to disease following vaccination but not infection ($V_{\sigma,a}$): | |
| 1mo | $\frac{dV_{\sigma,a}}{dt}=\left( S_{a-1,t}+M_{a-1,t}+R_{a-1,t} \right)\rho_{\sigma}-V_{\sigma,a,t}\left( w+\alpha_{a}+d_{a} \right)$ |
| 2-6mo | $\frac{dV_{\sigma,a}}{dt}=V_{\sigma,a-1,t}\alpha_{a-1}\left( \rho_{u}+\rho_{\sigma} \right)+\left( S_{a-1,t}+M_{a-1,t}+R_{a-1,t} \right)\rho_{\sigma}-V_{\sigma,a,t}\left( w+\alpha_{a}+d_{a} \right)$ |
| older ages | $\frac{dV_{\sigma,a}}{dt}=V_{\sigma,a-1,t}\alpha_{a-1}\left( \rho_{u}+\rho_{\sigma} \right)+\left( S_{a-1,t}+R_{a-1,t} \right)\rho_{\sigma}-V_{\sigma,a,t}\left( w+\alpha_{a}+d_{a} \right)$ |
| oldest age group | $\frac{dV_{\sigma,a}}{dt}=V_{\sigma,a-1,t}\alpha_{a-1}\left( \rho_{u}+\rho_{\sigma} \right)+\left( S_{a-1,t}+R_{a-1,t} \right)\rho_{\sigma}-V_{\sigma,a,t}\left( w+d_{a} \right)$ |
| Maternally immune ($M_{a}$): | |
| neonates | $\frac{dM_{a}}{dt}=b_{t}\left( 1-\frac{S_{15-44yo}}{N_{15-44yo}} \right)- M_{a,t}\left( \alpha_{a}+d_{a} \right)$ |
| 1-6mo | $\frac{dM_{a}}{dt}=M_{a-1,t}\alpha_{a-1}\rho_{u}-M_{a,t}\left( \alpha_{a}+d_{a} \right)$ |

† Where $\rho_{\lambda} = c_{t,a}\times f_{\lambda}$, $\rho_{\sigma} = c_{t,a}\times\left( 1 -f_{\lambda} \right)\times f_{\sigma}^{'}$, and $\rho_{u} = 1 - \rho_{\lambda} -\rho_{\sigma}$ to model vaccinations giving protection against infections, disease only, and vaccine failures combined with non-vaccination. Also, $f_{\sigma}^{'}=\frac{f_{\sigma} - f_{\lambda}}{1 - f_{\lambda}}$ to account for protection against breakthrough infections.

**Table S7**. Estimates of vaccine schedule effectiveness and waning used in the immunity tracking model.

| **Infectious disease / vaccine** | **Parameter†** | **Central estimate** | **Lower bound** | **Upper bound** | **Schedule and other specifications** | **Sources used** |
| --- | --- | --- | --- | --- | --- | --- |
| Diphtheria  (as part of pentavalent vaccine) | $f_{\lambda}$ | 0.600 | 0.510 | 0.680 | 3 infant doses at months 2, 4, and 6; booster around year 6. | [23, 24] |
|  | $f_{\sigma}$ | 0.810 | 0.740 | 0.860 |  | [24] |
|  | $w$ | 0.007 | 0.004 | 0.009 | After priming doses. Truelove et al. [24] also provide a low rate of waning. | [25] |
| Pertussis  (whole-cell, as part of pentavalent vaccine) | $f_{\lambda}$ | 0.830 | 0.600 | 0.920 | 3 infant doses at months 2, 4, and 6. Includes effect on infectiousness to others. | [[26]](https://www.sciencedirect.com/science/article/abs/pii/S0264410X03000070) |
|  | $f_{\sigma}$ | 0.940 | 0.880 | 0.970 | Effectiveness in the first 5 years of life against WHO definition of severe pertussis. | [[27, 28]](https://doi.org/10.1093/cid/ciw051) |
|  | $w$ | 0.043 | 0.034 | 0.059 | After priming doses. | [29] |
| Measles  (as part of MMR) | $f_{\lambda}$ | 0.850 | 0.250 | 0.970 | Dose 1 at 12mths, dose 2 at 18mths. | [30] |
|  | $f_{\sigma}$ | 0.941 | 0.883 | 0.983 |  | [31] |
|  | $w$ | 0.009 | 0.005 | 0.016 | After priming doses. Based on antibody waning. | [32] |
| Polio wild-type 1 or 3 / IPV-OPV sequential | $f_{\lambda}$ | 0.817 | 0.387 | 0.940 | IPV: 2 infant doses at months 1 and 2; OPV: 3 infant doses at months 2, 4, and 6 with boosters at 18mths and around year 6.  Based on faecal shedding after OPV challenge. | [33, 34] |
|  | $f_{\sigma}$ | 0.800 | 0.750 | 0.950 | Paralytic poliomyelitis. | [35, 36] |
|  | $w$ | 0.050 | 0.033 | 0.067 | After priming doses. Very limited data; made assumptions based on cited studies. | [37–39] |
| Polio vaccine-derived cVPD2 / IPV-OPV sequential | $f_{\lambda}$ | 0.220 | 0.180 | 0.422 | Shedding after mOPV2 challenge as the proxy. | [[40, 41]](https://doi.org/10.1093/cid/ciy633) |
|  | $f_{\sigma}$ | 0.800 | 0.750 | 0.950 | Paralytic poliomyelitis. | [35, 36] |
|  | $w$ | 0.050 | 0.033 | 0.067 | After priming doses. Very limited data; made assumptions based on cited studies. | [37–39] |
| *Haemophilus influenzae* type B  (as part of pentavalent vaccine) | $f_{\lambda}$ | 0.600 | 0.500 | 0.800 | 3 infant doses at months 2, 4, and 6. | [42, 43] |
|  | $f_{\sigma}$ | 0.180 | -0.020 | 0.330 | Radiologically confirmed pneumonia. | [44] |
|  | $f_{\sigma}$ | 0.940 | 0.780 | 0.980 | Meningitis. | [45, 46] |
|  | $w$ | 0.067 | 0.050 | 0.100 | After priming doses. Very limited data; made assumptions based on cited studies. | [47–49] |
| Pneumococcal conjugate vaccine  (10-valent) | $f_{\lambda}$ | 0.560 | 0.410 | 0.720 | 2 infant doses at months 2, 4; dose 3 at month 12. Protection against carriage among toddlers. | [50, 51] |
|  | $f_{\sigma}$ | 0.350 | 0.260 | 0.430 | Radiologically confirmed pneumonia. | [52, 53] |
|  | $f_{\sigma}$ | 0.920 | 0.580 | 0.990 | Meningitis, other invasive disease. | [54] |
|  | $w$ | 0.120 | 0.090 | 0.200 | Based on schedule with a booster in year 2. Waning may be faster without a booster [55]. | [[55, 56]](https://doi.org/10.1016/j.vaccine.2017.07.019) |
| Rotavirus (1-valent), ROTAVAC | $f_{\lambda}$ | 0.390 | 0.160 | 0.570 | 3 infant doses at months 2, 4, and 6.  Based on household transmission studies. | [57] |
|  | $f_{\sigma}$ | 0.889 | 0.809 | 0.935 | Hospitalisations due to rotavirus (‘developed country’ model). | [58, 59] |
|  | $w$ | 0.285 | 0.000 | 0.494 | After priming doses. | [60, 61] |

† $f_{\lambda}$: vaccine effectiveness against infection. $f_{\sigma}$ : vaccine effectiveness against severe disease. $w$: waning rate per year.

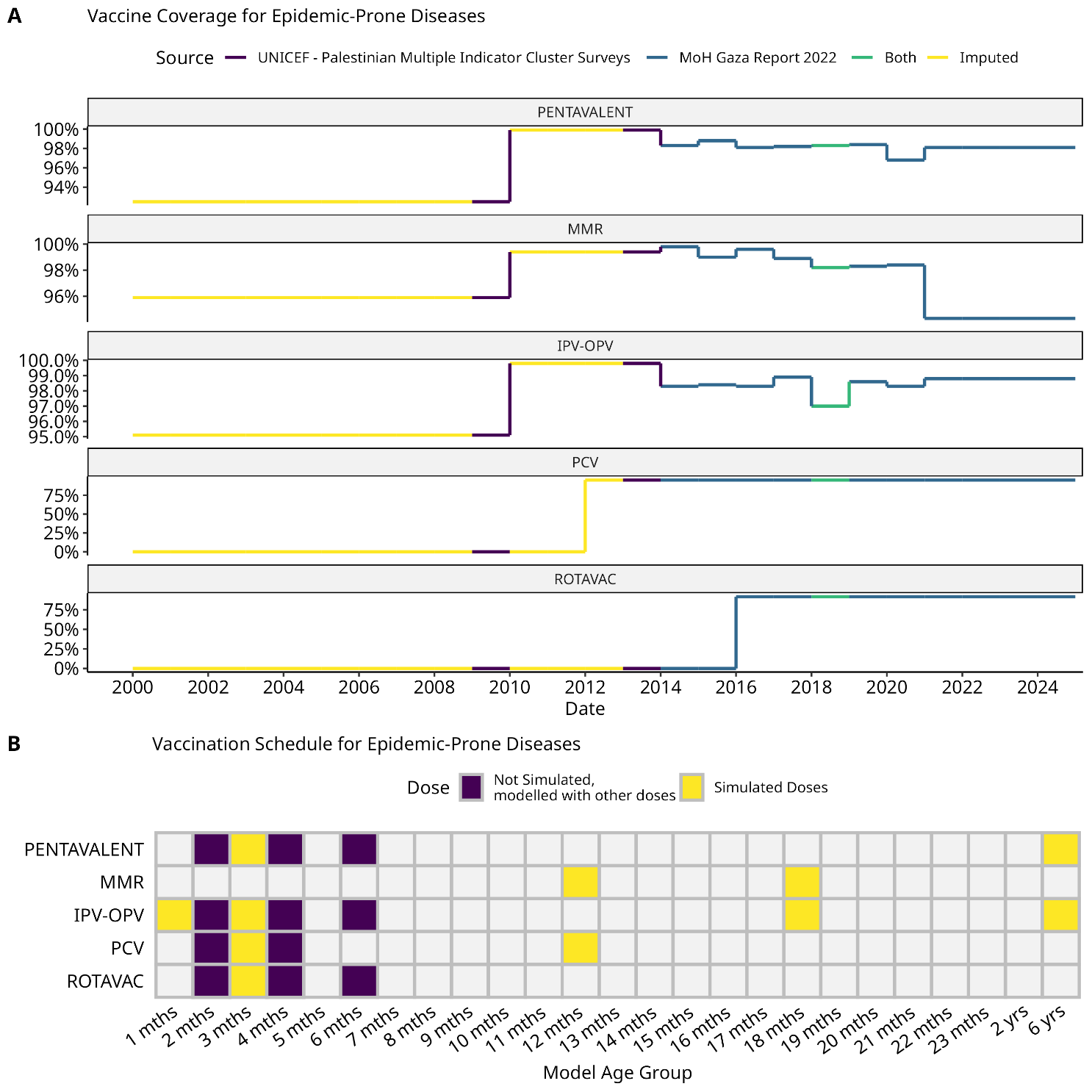

**Figure S3: Panel A:** Vaccine coverage values used in the immunity model, 2000-2025. Values taken from MoH Gaza Report 2022 [1] and UNICEF Palestinian Multiple Indicator Cluster Surveys (MICSs) 2010 [1], 2014 [1], and 2018 [1]. Where there were two alternative data sources, these were averaged. Missing values are imputed as the next available known value. Back-calculated values for 2000 are similar to coverage reported in MICS 2000 [1]. **Panel B:** Vaccine schedules used in the immunity model. For simplicity, doses between 2 months and 6 months are modelled as all occurring during month 3, polio birth doses during month 1 and booster doses at 12 months, 18 months and 6 years.

**Table S8**. Baseline susceptibility assumptions made for infectious diseases not included in the vaccination schedule.

| **Category** | **Infectious disease** | **Assumed % immune against infection and disease** | **Justification** |
| --- | --- | --- | --- |
| **Endemic** | Other bacterial gastroenteritis | No assumption | We assumed that the crisis had not yet resulted in reductions in immunity to these pathogens. |
|  | COVID-19 |  |  |
|  | Influenza, para-influenza |  |  |
|  | Other viral gastroenteritis |  |  |
|  | Respiratory syncytial virus |  |  |
| **Epidemic-prone** | Bacterial dysentery (*Shigella dysenteriae* type 1) | 0% | Likely to be circulating in Gaza based on non-zero reporting of shigellosis cases by the MoH pre-war, but at a low level. |
|  | Cholera | 0% | No known circulation in Gaza during the past decade. |
|  | Hepatitis A | neonates (0mo): 80%  1 to 11mo: 60%  12 to 59mo: 80%  5 to 9yo: 93%  older ages: 95% | Initial maternally acquired immunity followed by rapid, near-universal exposure in childhood. Values approximated from seroprevalence studies in Palestine [62] and from nearby countries [63]. |
|  | Hepatitis E | 0% | No known circulation in Gaza during the past decade. |
|  | Meningococcal meningitis | 0% | While *N. meningitidis* does circulate in Gaza, it is likely based on mortality data that transmission intensity is low. |
|  | Typhoid fever | 0% | Low level of transmission in previous years (≈ 15 cases reported per year between 2016 and 2022). |

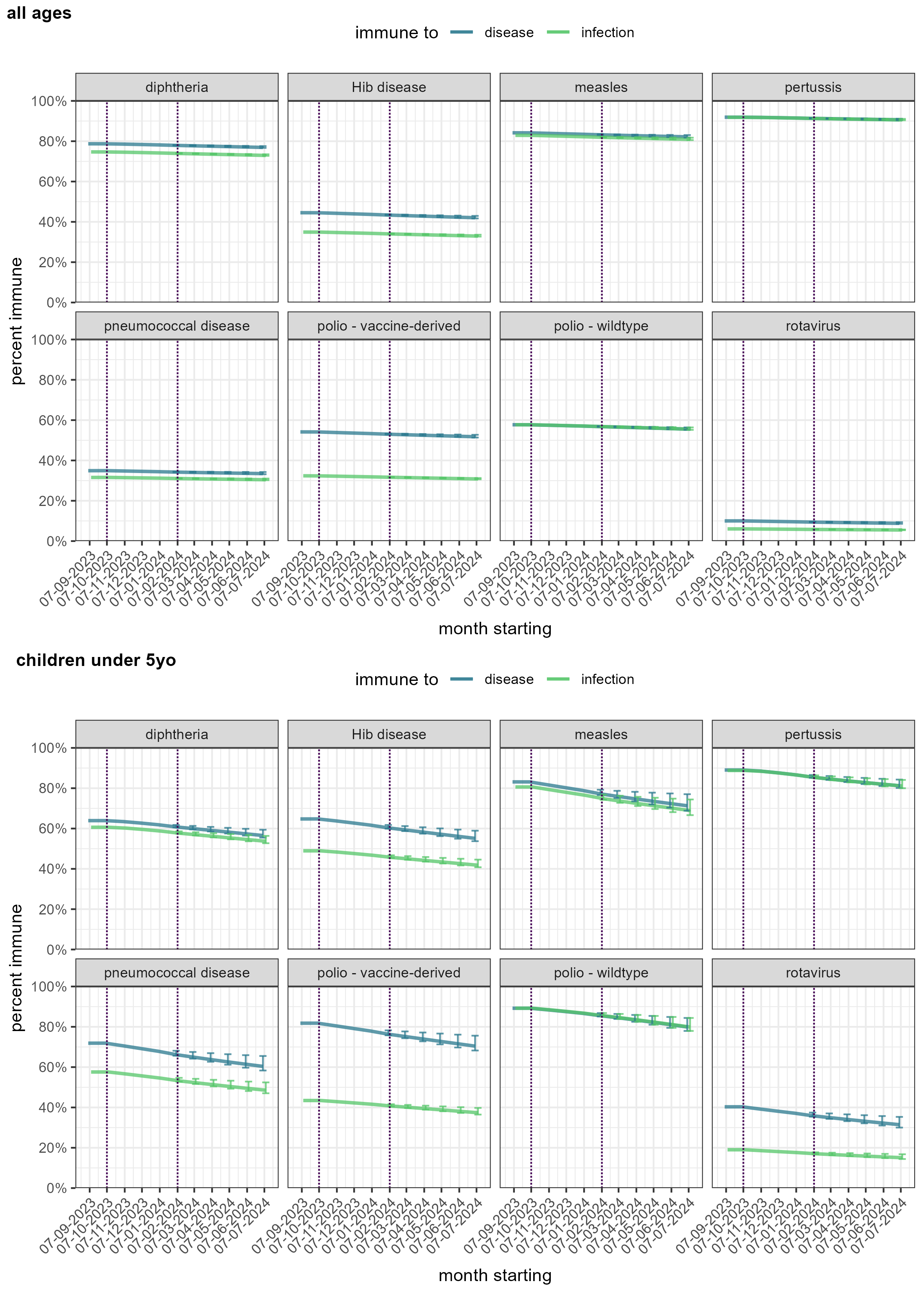

**Figure S4.** Estimated proportion of people with immunity to infection or disease, by month, age group and disease. The main line indicates the status quo scenario: vertical bars show the range from ceasefire (top) to escalation (bottom) scenarios.

#### Transmissibility, case-fatality and epidemic probability

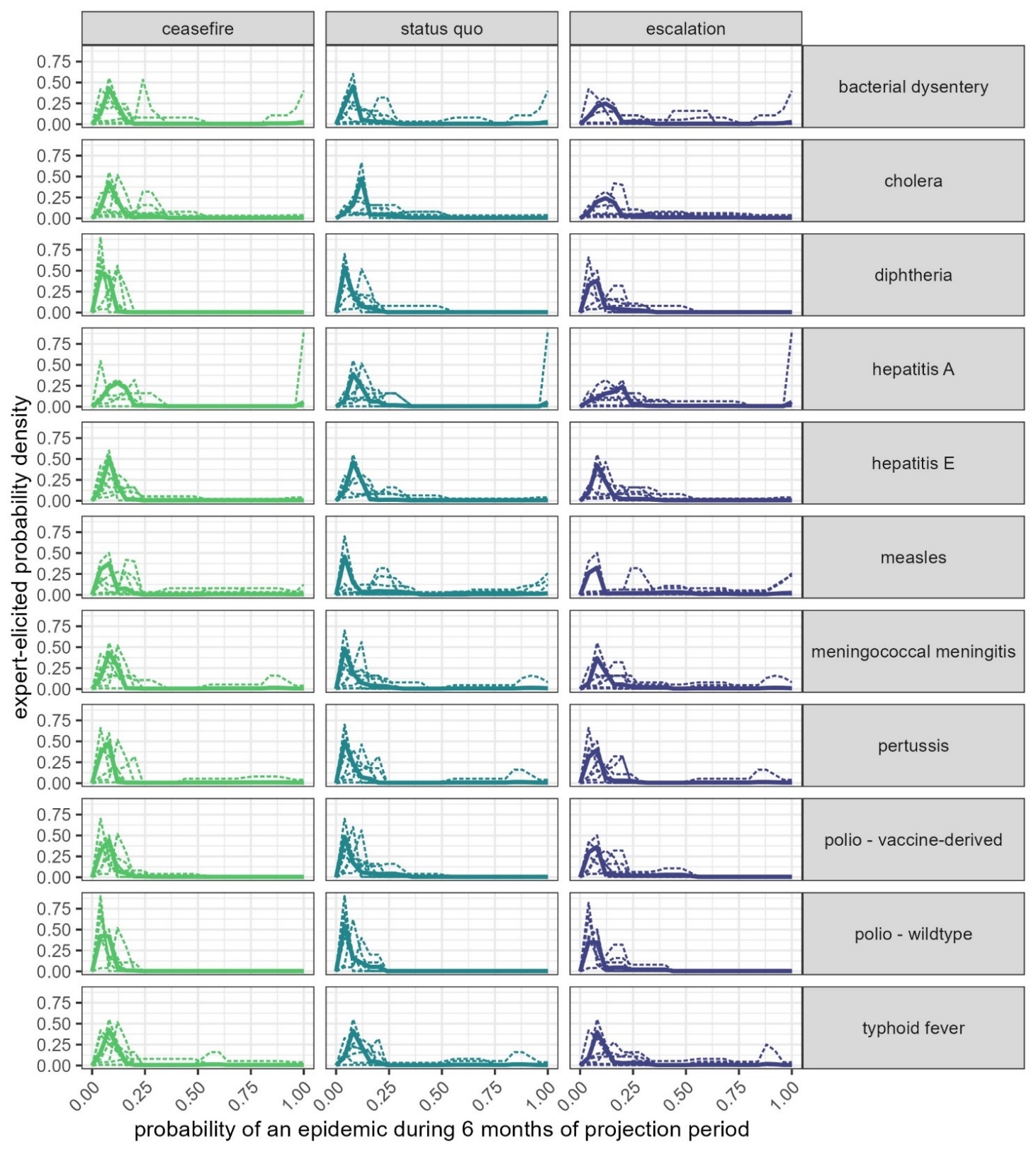

**Figure S5.** Expert-elicited probability of an epidemic in the next 6 months, by pathogen and scenario. Dotted lines indicate individual experts’ estimates. The thick line is the weighted mean value, used as model input.

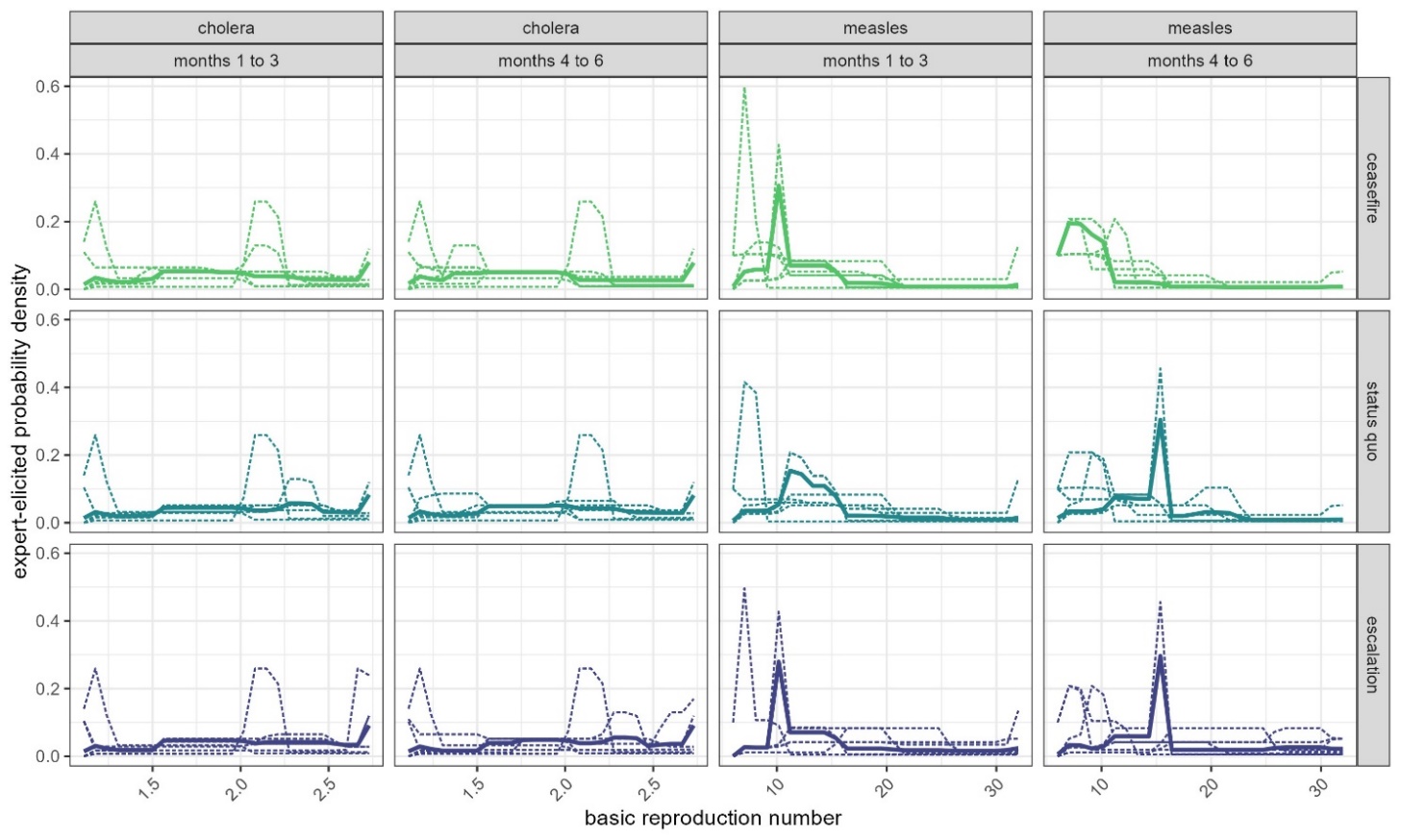

**Figure S6.** Expert-elicited distributions of the $\mathcal{R}_{0}$ of cholera and measles, by scenario. Dotted lines indicate individual estimates. The thick line is the weighted mean. Experts gave estimates for the first and second trimester of the projection period, as scenario values differed across these two sub-periods.

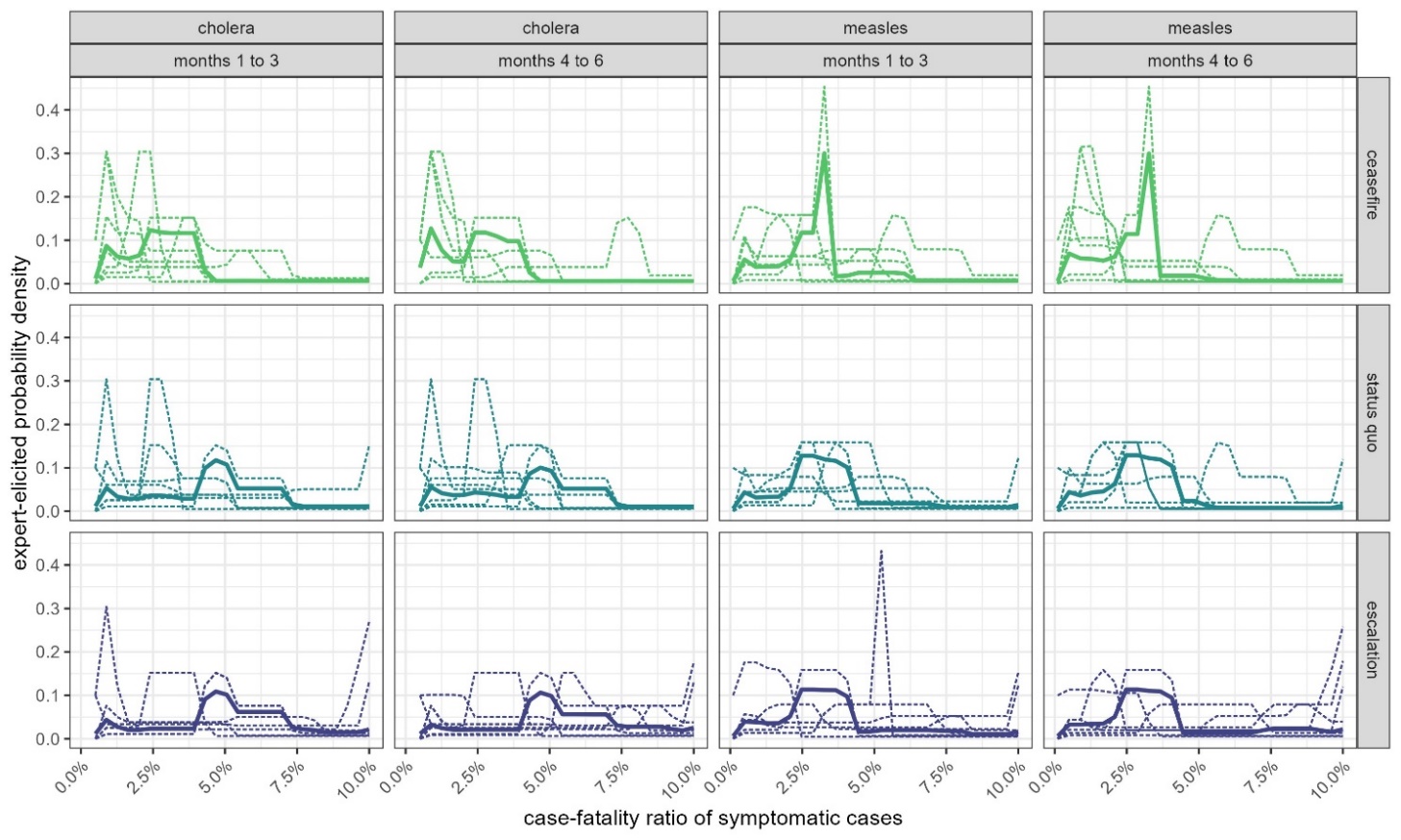

**Figure S7.** Expert-elicited distributions of the CFR of cholera and measles, by scenario. Dotted lines indicate individual estimates. The thick line is the weighted mean. Experts gave estimates for the first and second trimester of the projection period, as scenario values differed across these two sub-periods.

**Table S9.** Occurrence and timing of epidemics in past crises. Shaded cells are those for which the given pathogen caused an epidemic. Numbers in these cells indicate the recognised start of the epidemic, relative to the crisis’ start (in months).

| **Crisis typology** | **Crisis-affected region (date of major escalation or displacement)** | **Diphtheria** | **Measles** | **Pertussis** | **Meningo-coccal meningitis** | **Polio** | **Hepatitis A** | **Hepatitis E** | **Cholera** | **Typhoid fever** | **Bacterial dysentery** |
| --- | --- | --- | --- | --- | --- | --- | --- | --- | --- | --- | --- |
| Armed conflict | Iraq (Mar 2003) |  | 9 |  |  |  |  |  | 1 |  |  |
| Armed conflict | Darfur region, Sudan and Darfurian refugees to Chad (Dec 2003) |  | 4 |  | 13 | 6 |  | 5 | 32 |  | 7 |
| Earthquake | Kerman province, Iran (Dec 2003) |  |  |  |  |  |  |  |  |  |  |
| Armed conflict | North, South Kivu provinces, DRC (Jun 2004) |  |  |  |  |  |  |  | 6 |  |  |
| Flooding disaster (Indian Ocean tsunami) | Aceh province, Indonesia (Dec 2004) |  | 1 |  |  |  |  |  |  |  |  |
| Earthquake | Azad Jammu and Kashmir province, Pakistan (Oct 2005) |  |  |  |  |  |  |  |  |  |  |
| Armed conflict | Iraq (Feb 2006) |  | 36 |  |  |  |  |  | 17 |  |  |
| Armed conflict | Northern Province, Sri Lanka (Jan 2008) |  |  |  |  |  | 15 |  |  |  |  |
| Flooding disaster (Cyclone Nargis) | Ayeyarwady Region, Myanmar (Apr 2008) |  |  |  |  |  |  |  |  |  |  |
| Earthquake | Haiti (Jan 2010) |  |  |  |  |  |  |  | 9 | 2 |  |
| Drought | South-Central regions, Somalia and Somali refugees to Ethiopia, Kenya (Oct 2010) |  | 6 |  |  | 32 |  |  | 5 |  |  |
| Armed conflict | Ivory Coast (Nov 2010) |  | 2 |  | 14 |  |  |  | 18 |  |  |
| Armed conflict | Libya (Feb 2011) |  |  |  |  |  |  |  |  |  |  |
| Armed conflict | Syria and Syrian refugees to neighbouring countries (Mar 2011) |  | 12 |  |  | 31 |  |  |  | 23 |  |
| Armed conflict | South Kordofan, Blue Nile states, Sudan and refugees to South Sudan (Jun 2011) |  |  |  |  |  | 24 | 13 |  |  |  |
| Armed conflict | Tombouctou, Gao, Kidal, Taoudenit, Menaka regions, Mali (Jan 2012) |  |  |  |  |  |  |  | 6 |  |  |
| Armed conflict | Central African Republic (Dec 2012) |  | 1 |  |  |  |  |  | 36 |  |  |
| Flooding disaster (Typhoon Hayan) | Eastern Visayas (Region VIII), Philippines (Nov 2013) |  |  |  |  |  |  |  |  |  |  |
| Armed conflict | South Sudan (Dec 2013) |  | 10 |  |  | 11 |  | 20 | 1 |  |  |
| Armed conflict | Donetsk, Luhansk oblast, Ukraine (Apr 2014) |  |  |  |  |  |  |  |  |  |  |
| Armed conflict | Libya (May 2014) |  |  |  |  |  |  |  |  |  |  |
| Armed conflict | Yemen (Mar 2015) | 29 | 10 |  |  |  |  |  | 19 |  |  |
| Armed conflict | Borno, Adamawa, Yobe states, Nigeria (Jan 2015) |  |  |  | 24 | 19 |  | 29 | 2 |  |  |
| Armed conflict | Rohingya refugees in Cox Bazar camps, Bangladesh (Aug 2017) | 4 | 1 |  |  |  | 2 |  | 26 |  |  |
| Drought | South-Central regions, Somalia (Jan 2017) |  | 2 | 5 |  |  |  |  | 11 |  |  |
| Armed conflict | Northwest, Southwest regions, Cameroon (Sep 2017) |  |  |  |  |  |  |  | 8 |  |  |
| Armed conflict | Tigray region, Ethiopia and Tigrayan refugees to Sudan (Nov 2020) |  | 10 |  |  |  |  | 7 | 35 |  |  |
| Armed conflict | Nagorno-Karabakh region, Azerbaijan (Sep 2020) |  |  |  |  |  |  |  |  |  |  |
| **Totals** | Number of epidemics during the first 36mths | 2 | 13 | 1 | 3 | 5 | 3 | 5 | 16 | 2 | 1 |
|  | Percentage of crises with epidemics during the first 36mths | 7% | 46% | 4% | 11% | 18% | 11% | 18% | 57% | 7% | 4% |
|  | Median delay (months) between crisis onset and epidemic | 17 | 6 | 5 | 14 | 19 | 15 | 13 | 10 | 13 | 7 |

Sources: Warsame et al. [64], complemented by searches of the World Health Organization’s Disease Outbreak News archive (<https://www.who.int/emergencies/disease-outbreak-news>) and keyword (‘[disease name] OR [pathogen name]’ + ‘outbreak OR epidemic’) searches of PubMed/Medline.

**Table S10.** Values of the basic reproduction number, for each epidemic-prone infection.

| **Infectious disease** | **Minimum** | | | **Maximum** | | |
| --- | --- | --- | --- | --- | --- | --- |
|  | value | source | notes | value | source | notes |
| bacterial dysentery (*S. dysenteriae* type 1) | 1.10 | [65] | US outbreaks, but of *S. sonnei* | 2.20 | [65] | US outbreaks, but of *S. sonnei* |
| cholera | 1.11 | [66] | Zimbabwe | 2.73 | [66] | Zimbabwe |
| diphtheria | 1.70 | [24] | Synthesis of various studies | 7.10 | [[67] [68]](https://bmcmedicine.biomedcentral.com/articles/10.1186/s12916-019-1288-7) | Mean of two refugee camps, Bangladesh |
| hepatitis A | 1.10 | [69] | USA | 2.70 | [70] | children in China |
| hepatitis E | 2.11 | [71] | Uganda | 8.50 | [72] | Refugees in Uganda |
| measles | 6.00 | [73] | Systematic review of outbreaks | 32.00 | [73] | outbreaks |
| meningococcal meningitis | 1.31 | [74] | Italy | 2.50 | [75] | Nigeria |
| pertussis | 5.50 | [76] | Europe | 17.00 | [77] | Systematic review |
| polio - vaccine-derived | 1.62 | [78] | Latest polio outbreak in Israel | 12.00 | [79] | Afghanistan, Pakistan |
| polio - wildtype | 1.62 | [78] | Latest polio outbreak in Israel | 12.00 | [79] | Fully reverted vdPV seems to have similar transmissibility as wPV [80]. |
| typhoid fever | 2.80 | [81] | no variability found in the literature | 2.80 | [81] | no variability found in the literature |

**Table S11.** Estimates of case fatality ratio of symptomatic cases, by age and by pathogen.

| **Infectious disease** | **Case-fatality ratio, age 12-59mo (%)** | | | | **Relative risk, compared to the 12-59mo category** | | | | | | | | | | | | | **Source** |
| --- | --- | --- | --- | --- | --- | --- | --- | --- | --- | --- | --- | --- | --- | --- | --- | --- | --- | --- |
|  | minimum  (source) | | maximum  (source) | | <1  mo | 1-11  mo | *12-59 mo* | 5-9  yo | 10-14  yo | 15-19  yo | 20-29  yo | 30-39  yo | 40-49  yo | 50-59  yo | 60-69  yo | 70-79  yo | ≥80  yo |  |
| bacterial dysentery | 0.6 | [82] | 7.4 | [82] | 1.72 | 1.72 | *1* | 0.69 | 0.69 | 0.44 | 0.44 | 0.44 | 0.44 | 0.44 | 0.44 | 0.44 | 0.44 | [83] |
| cholera | 0.5 | [84] | 10.0 | [84] | 1.00 | 1.00 | *1* | 1.00 | 1.00 | 1.00 | 1.00 | 1.00 | 1.80 | 1.80 | 3.00 | 3.00 | 3.00 | [85] |
| diphtheria | 5.0 | [24] | 50.0 | [24] | 1.00 | 1.00 | *1* | 0.34 | 0.34 | 0.34 | 0.42 | 0.42 | 0.42 | 0.42 | 0.42 | 0.42 | 0.42 | [24] |
| hepatitis A | 0.02 | [86] | 1.8 | [87] | 1.00 | 1.00 | *1* | 1.00 | 1.00 | 1.00 | 1.35 | 1.35 | 7.89 | 7.89 | 14.88 | 14.88 | 14.88 | [88] |
| hepatitis E | 1.6 | [89] | 6.8 | [89] | 1.00 | 1.00 | *1* | 1.00 | 1.00 | 1.00 | 1.00 | 1.00 | 1.00 | 1.00 | 1.00 | 1.00 | 1.00 | [90, 91] |
| measles | 0.1 | [92] | 10.0 | [92] | 1.00 | 1.00 | *1* | 0.26 | 0.26 | 0.26 | 0.26 | 0.26 | 0.26 | 0.26 | 0.26 | 0.26 | 0.26 | [92] |
| meningococcal meningitis | 0.0 | [93] | 50.0 | [93] | 1.00 | 1.00 | *1* | 1.00 | 1.00 | 1.00 | 1.00 | 1.00 | 1.00 | 1.00 | 1.00 | 1.00 | 1.00 | [93] |
| pertussis | 0.7 | [94] | 15.0 | [94] | 3.70 | 3.70 | *1* | 0.10 | 0.10 | 0.10 | 0.10 | 0.10 | 0.10 | 0.10 | 0.10 | 0.10 | 0.10 | [95] |
| polio - vaccine-derived | 6.0 | [96] | 40.0 | [96] | 0.90 | 0.90 | *1* | 2.00 | 2.20 | 3.60 | 3.60 | 4.70 | 4.70 | 4.70 | 4.70 | 4.70 | 4.70 | [97] |
| polio - wildtype | 6.0 | [96] | 40.0 | [96] | 0.90 | 0.90 | *1* | 2.00 | 2.20 | 3.60 | 3.60 | 4.70 | 4.70 | 4.70 | 4.70 | 4.70 | 4.70 | [97] |
| typhoid fever | 0.6 | [98] | 8.9 | [98] | 1.00 | 1.00 | *1* | 1.00 | 1.00 | 1.00 | 1.00 | 1.00 | 1.00 | 1.00 | 1.00 | 1.00 | 1.00 | [98] |

### References

Automatic citation updates are disabled. To see the bibliography, click Refresh in the Zotero tab.
